# Establishment of an Anti-Nephrin Autoantibody Assay and the Predictive and Diagnostic Significance of Pre-treatment Antibody Titers in Pediatric Nephrotic Syndrome

**DOI:** 10.64898/2026.10.24.26366391

**Authors:** Tomoko Horinouchi, Masaya Muto, Takaya Ozeki, China Nagano, Yuta Inoki, Yuta Ichikawa, Nana Sakakibara, Maria Kiriyama, Yumiko T. Sakai, Eri Morita Yamamoto, Valentina Podolinnaia, JeeEun Kim, Aoi Tanaka, Yusuke Atarashi, Nanami Nagatsuka, Keiko Miwa, Mami Onishi, Akshay Ganguly, Kazuma Kita, Bishnu Devi Maharjan, Tomohiko Yamamura, Koichi Kamei, Riku Hamada, Natsumi Yamamura, Miwa Goto, Hideki Ban, Yoshitsugu Kaku, Rie Kuroki, Naoya Fujita, Hiroki Miyano, Tomoo Kise, Rika Fujimaru, Hiroshi Kaito, Erika Hidawa, Yohei Ikezumi, Wataru Shimabukuro, Yusuke Okuda, Yuko Shima, Taishi Nada, Shin’ichi Akiyama, Kazumichi Fujioka, Masanori Miyanishi, Kenji Ishikura, Koichi Nakanishi, Yoshitaka Isaka, Shoichi Maruyama, Amane Harada, Kandai Nozu, Kazumoto Iijima

## Abstract

**Introduction:** Anti-nephrin autoantibodies have been implicated in podocytopathies, including minimal change disease and focal segmental glomerulosclerosis. However, their prevalence, longitudinal dynamics, and clinical significance in first-onset nephrotic syndrome (NS) including steroid-sensitive NS (SSNS) and steroid-resistant NS (SRNS) remain unclear.

**Methods:** We established a highly sensitive anti-nephrin autoantibody assay using a modified recombinant nephrin antigen and the automated immunoassay platform, and applied it to a prospective Japanese pediatric, multicenter, cohort with first-onset NS and an independent validation cohort. Anti-nephrin autoantibody titers were evaluated before and after treatment and analyzed in relation to clinical phenotypes, genetic risk alleles, and tissue findings. The assay was validated using immunoprecipitation– Western blotting (IP-WB) in representative samples, assessing IgG-nephrin colocalization in evaluable kidney biopsy specimens, and antigen inhibition assays in all antibody-positive samples from the validation cohort.

**Results:** In the pediatric discovery cohort (n = 103), all patients with SSNS (n = 83, 100%) and nine of 16 with SRNS (56.3%) were positive for anti-nephrin autoantibodies, whereas all patients with IgA nephropathy (IgAN) with NS onset were negative (n = 4, 0%). Standard-range positivity was associated with SSNS (77/77, 100%), high-titer positivity with SRNS (9/15, 60.0%), and antibody negativity was restricted to non-SSNS diseases, comprising SRNS (7/11, 63.6%) and IgAN (4/11, 36.4%) (*P* < 0.0001). Similarly, in the independent validation cohort (n = 42), high-titer positivity was associated with SRNS (1/1, 100%), standard-range positivity with SSNS (30/33, 90.9%), and antibody negativity identified IgAN and membranous nephropathy (5/8, 62.5%) or SRNS (3/8, 37.5%). Orthogonal validation by IP-WB, IgG-nephrin colocalization, and antigen inhibition assays was concordant.

**Conclusions:** Anti-nephrin autoimmunity is a hallmark of NS and identifies distinct clinical phenotypes at disease onset. The automated immunoassay for anti-nephrin autoantibodies enables rapid, highly sensitive, and automated antibody measurement and may serve as a diagnostic triage tool for early etiologic stratification and clinical characterization of NS at disease onset.

**Lay Summary:** Nephrotic syndrome is a kidney disease in which proteins leak into the urine because of a damaged kidney filtration barrier. Recent studies suggest that some patients produce autoantibodies against nephrin, a key protein required for normal kidney filtration. However, accurately measuring these antibodies has been difficult. In this study, we developed a highly sensitive automated blood test for anti-nephrin autoantibodies and used it in Japanese children with newly diagnosed nephrotic syndrome. We found that all children with steroid-sensitive nephrotic syndrome were positive for anti-nephrin autoantibodies at disease onset. In contrast, children with very high antibody levels or no detectable antibodies were more likely to have steroid-resistant disease or other kidney diseases. These findings were confirmed in an independent validation cohort. This blood test may help improve diagnosis, predict treatment response, and support more personalized management of nephrotic syndrome.

## Introduction

Nephrotic syndrome (NS) in children is clinically and pathophysiologically heterogeneous, encompassing steroid-sensitive (SSNS) and steroid-resistant (SRNS) forms.^1^ Most children achieve remission with corticosteroid therapy; however, some do not and develop SRNS, which is associated with a high risk of progression to chronic kidney disease.^2^ The underlying mechanisms remain incompletely understood, especially in first-onset cases without identifiable monogenic defects.

Nephrin, encoded by *NPHS1* gene, is a critical structural protein of the podocyte slit diaphragm, essential in maintaining the glomerular filtration barrier.^3^ Pathogenic *NPHS1* variants cause congenital NS of the Finnish type.^3^ Moreover, genome-wide association studies of SSNS have identified several susceptibility genes, including loci within the *HLA* region, and risk alleles in *NPHS1* have been reported.^4–10^ These *NPHS1* risk alleles are specific to East Asian populations.^8, 9^ Beyond genetic defects, accumulating evidence suggests that nephrin can also become the target of pathogenic autoantibodies. Anti-nephrin autoantibodies have been identified in a subset of patients with minimal change disease and focal segmental glomerulosclerosis.^11–16^

However, several important issues remain unresolved. The prevalence of anti-nephrin autoantibodies in pediatric patients with first-onset NS and the longitudinal changes in their titers before and after treatment have not been systematically examined, and differences between SSNS and SRNS remain insufficiently characterized. Moreover, the relationship between anti-nephrin autoantibody titers and genetic background, including *NPHS1* variants and *HLA* risk alleles, has not been adequately investigated.

Furthermore, anti-nephrin autoantibody detection remains technically challenging, and its prevalence varies.^17, 18^ This discrepancy arises because circulating autoantibodies are of extremely low titer, making their detection highly susceptible to the timing of sample collection (i.e., the patient’s treatment status) and the sensitivity of the assay system. To overcome these limitations, various highly sensitive and specific detection strategies have been developed, including signal-enhanced enzyme-linked immunosorbent assay (ELISA),^11, 19, 20^ immunoprecipitation (IP) followed by Western blotting (WB),^11, 12^ and a hybrid IP–ELISA quantification approach.^12^ However, many of these assay systems inevitably sacrifice throughput to achieve sufficient sensitivity, and differences in glycosylation patterns arising from the source of recombinant nephrin can lead to substantial background noise and false-positive signal.^18, 21^ Moreover, the nephrin antigen contains multiple free cysteines within its extracellular domain region,^22^ which are generally known to negatively impact protein stability by inducing aggregation through unwanted intermolecular disulfide bonding.^23–25^ Consequently, this structural instability poses a significant risk of compromising antigen immunoreactivity and overall assay performance.^17^ Given the inconsistent prevalence in previous reports, the trade-off between sensitivity and assay throughput, and the structural variations on the antigen side, careful assay validation, appropriate controls, and cross-validation using complementary methodologies are essential before anti-nephrin autoantibodies can be reliably implemented as clinical biomarkers or used to guide therapeutic decision-making.^17, 18, 26^

To address these gaps, we established a highly sensitive and high-throughput automated immunoassay system using a modified nephrin antigen and applied it to a well-characterized, multicenter, Japanese pediatric cohort with first-onset NS. We compared pre- and post-treatment titers, evaluated associations with disease phenotype (SSNS, SRNS), genetic background, and biopsy findings.

## Methods

### Patients

#### Pediatric idiopathic NS cohort (discovery)

We enrolled children aged ≥1 year and <15 years who developed first-onset NS between July 2024 and December 2025 at 29 participating medical institutions across Japan. NS was defined as serum albumin level ≤2.5 g/dL and urinary protein-to-creatinine ratio ≥2.0 g/g.^27^ Serum or plasma samples were collected before treatment initiation. Post-treatment samples were obtained within 35 days after treatment initiation, either after remission or during this period, if remission was not achieved.

#### Pediatric idiopathic NS cohort (validation)

The independent validation cohort included children aged ≥1 year and <15 years who developed first-onset NS between January 2026 and July 2026 at 24 participating medical institutions across Japan. Serum or plasma samples were collected before treatment initiation.

#### Control cohort

To establish assay-specific positivity thresholds, we included 30 pediatric controls who presented to a pediatric emergency clinic with acute self-limited illnesses, such as upper respiratory tract infections and gastroenteritis. To assess disease specificity, we additionally included pre-treatment biopsy-proven pediatric IgA nephropathy (IgAN) and IgA vasculitis nephritis (IgAVN) identified outside the NS cohorts, as well as 30 healthy adult controls.

#### Treatment and outcomes

All patients with pediatric idiopathic NS were treated according to the Japanese clinical practice guidelines for pediatric idiopathic NS,^27^ consistent with international guidelines.^28^ Initial therapy consisted of oral prednisolone administered at 60 mg/m²/day or 2 mg/kg/day (maximum 60 mg/day). SRNS was defined as failure to achieve complete remission after 4 weeks of treatment initiation. Clinical data were collected from the electronic medical records at each participating institution.

#### Generation of recombinant wild-type and mutant nephrin proteins

Recombinant human nephrin (extracellular domain, amino acids 23-1058) and free cysteine-substituted mutants were generated using the pcDNA3.4 vector. Hexa histidine (6xHis)-tagged proteins were transiently expressed in Expi293F cells and purified from culture supernatants by His-tag affinity chromatography. NanoDrop spectrophotometry and sodium dodecyl sulfate–polyacrylamide gel electrophoresis (SDS-PAGE) were used to assess protein concentration and purity, respectively. Supplementary Methods describe the detailed protocols.

#### Establishment of an automated immunoassay for anti-nephrin autoantibody detection

We established a chemiluminescent enzyme immunoassay using the HISCL-5000 system (Sysmex, Kobe, Japan) to quantify anti-nephrin autoantibodies. The assay was based on a two-step immunocapture principle using magnetic particles coated with recombinant human nephrin proteins. Serum or plasma samples were first diluted 40-fold with a 4-(2-hydroxyethyl)-1-piperazineethanesulfonic acid-based sample diluent and filtered through a 0.22-μm membrane. These pre-processed samples were incubated with the nephrin antigen-coated magnetic particles. Autoantibodies captured on the microparticles were detected using an alkaline phosphatase-fused anti-human IgG antibody (Sysmex, Kobe, Japan) and the chemiluminescent substrate CDP-Star (Applied Biosystems, Bedford, MA). Detailed descriptions of the reagents, optimization of the antigen, and the specific measurement procedures are provided in the Supplementary Methods.

The positivity threshold was established using serum samples from pediatric controls. During this evaluation, few of these control samples had non-specific signals due to non-specific adsorption of IgG. To eliminate these non-specific background signals, we adopted a subtraction approach to determine the ΔSU/mL value, wherein the signal measured using bovine serum albumin-coated magnetic particles was subtracted from the signal obtained using nephrin antigen-coated magnetic particles.

#### Analytical performance of the assay

We comprehensively validated the analytical performance of the automated immunoassay for anti-nephrin autoantibodies, evaluating the calibration curve, quantitative range, intra- and inter-assay reproducibility, sample stability, concordance between serum and plasma measurements, and potential interfering substances. Detailed experimental protocols are provided in the Supplementary Methods.

#### Detection of anti-nephrin autoantibodies by IP-WB

Representative positive and negative samples were evaluated using a previously reported IP-WB method to validate the automated immunoassay for circulating anti-nephrin autoantibodies.^11, 12^ Recombinant nephrin protein was incubated with patient serum or plasma samples and immunoprecipitated using protein G beads.

Immunoprecipitated complexes were separated by SDS-PAGE and analyzed by Western blotting using an anti-nephrin antibody. Detailed protocols are provided in the Supplementary Methods.

#### Immunohistological assay

Tissue staining to validate the circulating anti-nephrin autoantibody assay was performed in evaluable cases from the pediatric NS discovery cohort as previously reported.^29^ Double-immunofluorescence of nephrin and IgG was performed on frozen kidney sections using a premixed Alexa 546–anti-nephrin (Cat. No. 29050) (Immuno-Biological Laboratories, Fujioka, Japan) and Alexa 488–anti-IgG (not commercially available; Immuno-Biological Laboratories, Fujioka, Japan) antibody cocktail, with colocalization visualized as yellow fluorescence. Images were acquired using a KEYENCE BZX810 fluorescence microscope (KEYENCE Corporation, Osaka, Japan) with optical sectioning and analyzed with BZX800 software.

#### Antigen-inhibition assay

An antigen inhibition assay was performed to assess the specificity of anti-nephrin autoantibody reactivity. We incubated 9 μL of serum or plasma sample with 1 μL of recombinant human nephrin antigen solution at concentrations of 0 or 3,000 μg/mL, yielding final antigen concentrations of 0 or 300 μg/mL. After incubation at 37°C for 1 hour, the sample-antigen mixtures were diluted 40-fold with sample diluent, filtered, and analyzed immediately using the automated immunoassay platform according to the standard measurement protocol. A reduction in the anti-nephrin autoantibody signal following preincubation with nephrin antigen was interpreted as evidence of specific antigen-antibody binding.

#### Genotyping

In the pediatric idiopathic NS discovery cohort, genomic DNA was extracted from peripheral blood lymphocytes. Single-nucleotide polymorphisms (SNPs) in *HLA* locus (rs3129888; tag SNP for HLA-DRB1*08:02 and HLA-DRB1*08:02-DQB1*03:02 in the Japanese population ^5^) and *NPHS1* (rs2285450; synonymous variant) were assessed using SNP array analysis.^30^ *NPHS1* rs2285450 have shown genome-wide significant risk in Japanese, Korean, and South Asian populations, and their associations were further confirmed by trans-ethnic meta-analyses encompassing Japanese, Korean, South Asian, African, European, Hispanic, and Maghrebian populations.^8^ The *HLA* risk variant (rs3129888) was analyzed using a carrier-based model, whereas *NPHS1* risk variant (rs2285450) was analyzed according to genotype (non-carriers, heterozygotes, and homozygotes) to assess a potential allele dose effect. Targeted next-generation panel sequencing using a kidney disease gene panel was performed according to previously reported methods in patients who failed to achieve remission with multiple immunosuppressive agents.^31^

#### Ethical considerations

The study was conducted according to the principles of the Declaration of Helsinki and was approved by the Institutional Review Board of Kobe University and the participating institutions (approval numbers: B220129 and B210152). Written informed consent was obtained from participants or their legal guardians for prospective sample collection. For previously collected samples and clinical information, an opt-out consent procedure was implemented according to the ethical guidelines for medical and health research involving human subjects.

#### Statistics

Patient background characteristics are expressed as median (interquartile range) for continuous variables. Categorical variables were compared using contingency table analyses. Fisher’s exact test, including its extension for larger contingency tables when appropriate, was applied when expected cell counts were small, with corresponding odds ratio (OR) and 95% confidence interval (CI). Spearman’s rank correlation was used to assess correlations between pre-treatment anti-nephrin autoantibody titers and time to remission, and to evaluate concordance between antibody titers measured in paired serum and plasma samples. Wilcoxon matched-pairs signed-rank test was used for paired comparisons of anti-nephrin autoantibody titers before and after treatment. Comparisons of anti-nephrin autoantibody titers were performed using the Mann– Whitney U test for two-group comparisons and the Kruskal–Wallis test for three-group comparisons. Monotonic associations with *NPHS1* allele dosage were assessed using Spearman’s rank correlation. To determine the optimal cut-off value of the anti-nephrin autoantibody titer for differentiating SSNS from SRNS, a receiver operating characteristic (ROC) curve was constructed with corresponding sensitivity and specificity.

## Results

### Establishment and analytical validation of an automated immunoassay for anti-nephrin autoantibody detection

#### Optimization of the nephrin antigen for enhanced reactivity

To overcome the limitations of conventional assays regarding sensitivity and reliability, we optimized the antigen design by focusing on three free cysteine residues (Cys101, Cys721, and Cys974) within the nephrin molecule. We generated variants in which these residues were substituted with serine (SSS), alanine (AAA), or valine (VVV). Each variant was incorporated into an automated immunoassay, and assay reactivity was compared using serum samples from patients with NS. All substituted variants produced stronger antibody signals than the wild-type antigen (CCC), with the Val-substituted variant demonstrating the highest signal intensity (Figure 1a). Western blot analysis showed that substitution of the three free cysteine residues reduced the slower-migrating species observed in wild-type under non-reducing conditions (Supplementary Figure S1, Supplementary methods). These findings suggest that substitution of these free cysteine residues improved antigen homogeneity, thereby enhancing assay sensitivity. Further analysis of individual substitution sites revealed that mutations at Cys721 and Cys974 particularly contributed to the enhanced reactivity with anti-nephrin autoantibodies (Figure 1b).

**Figure 1.**
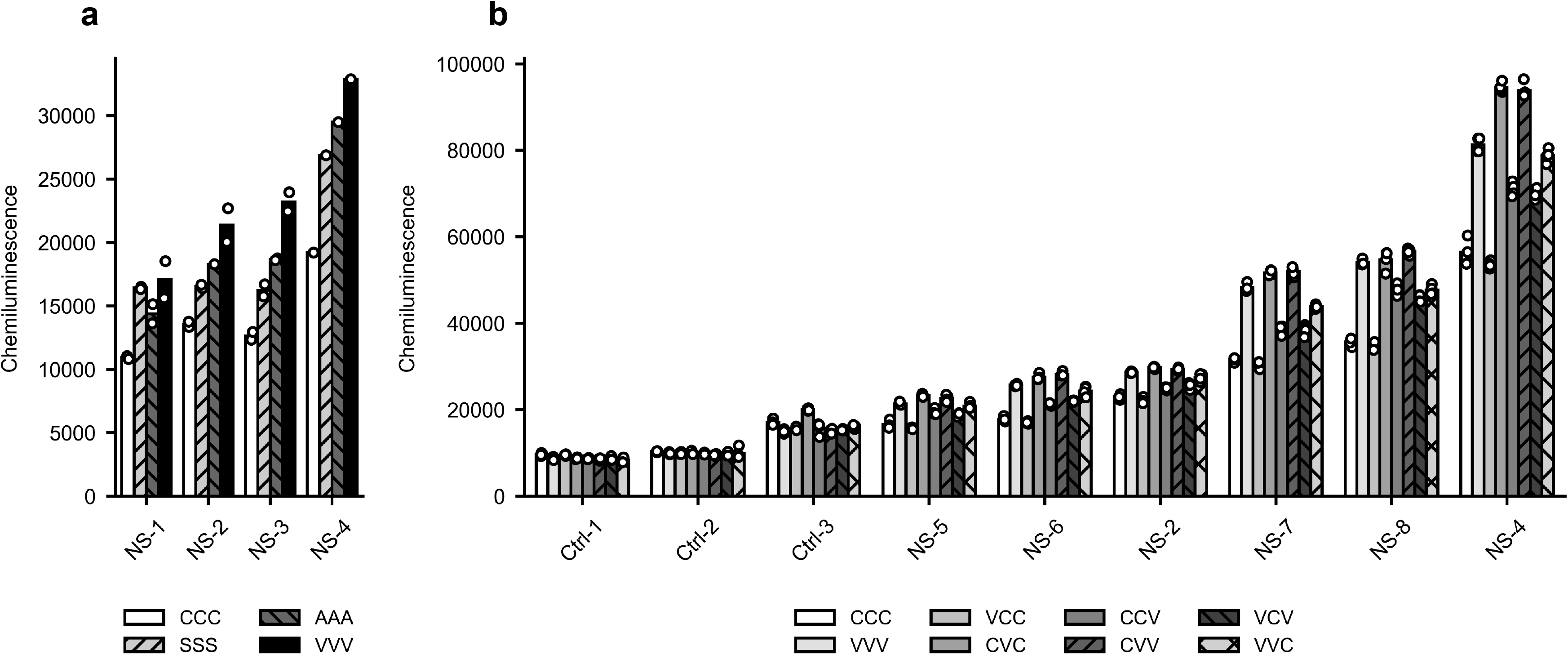
Optimization of the nephrin antigen for enhanced reactivity in the anti-nephrin antibody assay. **(a)** Immunoreactivities of wild-type (CCC) nephrin and its triple-mutated variants. Three free cysteine residues (Cys101, Cys721, and Cys974) were substituted with serine (SSS), alanine (AAA), or valine (VVV). Reactivity was evaluated using serum samples from four patients with nephrotic syndrome (NS). Bars represent the mean values, and dots indicate individual measurements (n = 2 for three samples; n = 1 for one sample). **(b)** Evaluation of individual substitution sites using CCC nephrin and single, double, or triple Val-substituted mutants. Reactivity was analyzed using samples from healthy controls (Ctrl-1, Ctrl-2, and Ctrl-3) and NS patients. The three-letter codes indicate the amino acids at positions 101, 721, and 974, respectively, from left to right (C: Cysteine; A: Alanine; V: Valine). Bars represent the mean values, and dots indicate individual measurements (n = 4 for all samples).

#### Analytical performance of the automated immunoassay

A calibration curve expressed in assay-specific units (SU/mL), was generated by gravimetric dilution of high-titer positive samples, with relative errors ranging from −4.9% to 3.2% (Supplementary Figure S2a, Supplementary Table S1). The assay demonstrated a wide quantitative range of 152 to 57,419 SU/mL based on the limit of quantitation (LoQ) and dilution linearity assessed using three high-titer NS samples (Supplementary Figure S1b, Supplementary Table S2). Precision was high, with intra-assay and inter-assay coefficients of variation levels were 1.1–1.7% and 3.1–5.4%, respectively, across three concentration levels (Supplementary Figure S2c, S2d, Supplementary Table S3). Antibody titers remained stable within 93.3% to 108.5% of baseline values under various storage conditions (−20°C for 7 days, and 4°C or 25°C for 1 day) and after up to five freeze-thaw cycles, with no significant fluctuations observed in stability testing (Supplementary Figure S1e, S1f). Furthermore, we assessed 20 paired serum and plasma samples from pediatric patients with NS (14 in the active phase and 6 in remission) to evaluate sample matrix compatibility. Antibody titers in serum and plasma showed an excellent correlation (Spearman’s ρ = 0.998), demonstrating that both sample types are equally suitable for high-precision evaluation (Supplementary Figure S2g). The assay showed no significant interference from major blood components, including free and conjugated bilirubin, hemolytic hemoglobin, chyle, and rheumatoid factor, even at the maximum concentrations specified by the commercial kits; the maximum deviation observed was −6.4% for chyle (Supplementary Table S4).

#### Validation of the Automated Immunoassay by IP-WB

Samples included pediatric anti-nephrin autoantibody-positive and -negative NS cases at disease onset and remission and healthy adult controls. Anti-nephrin autoantibody positivity determined by the automated immunoassay showed strong concordance with IP-WB. Representative immunoblots are shown in Supplementary Figure S3.

#### Comparison with renal tissue immunostaining

Detailed findings for the eight biopsied cases are presented in Supplementary Table S5 and Supplementary Figure S4. The status of IgG–nephrin colocalization was consistent with circulating anti-nephrin autoantibody titers. However, one case was considered inconclusive, potentially due to treatment-related modification of the pathological findings.

### Anti-nephrin autoantibody titers at disease onset and clinical characteristics in pediatric idiopathic NS

#### Pre-treatment anti-nephrin autoantibody titers and clinical outcomes in the pediatric discovery cohort

We enrolled 103 children with newly diagnosed NS during the study period. The clinical characteristics of the pediatric patients are shown in Supplementary Table S6. Ultimately, 83 patients were diagnosed with SSNS, 16 with SRNS, and 4 with glomerulonephritis (GN). All GN cases were diagnosed with IgAN.

The positivity threshold was established using serum samples from 30 pediatric acute disease controls. Following background correction, a single control sample exhibited an explicitly elevated signal, presumably derived from anti-nephrin autoantibodies. This sample was statistically identified as a clear outlier using Tukey’s fences method and was subsequently excluded. Among the remaining 29 control samples, the highest recorded value of 527 ΔSU/mL was established as the threshold for anti-nephrin autoantibody positivity (Figure 2, Supplementary Figure S5). All biopsy-proven IgAN (n = 8) and IgAVN (n = 11) cases as well as adult healthy controls (n = 30) were negative for anti-nephrin autoantibodies (Figure 2).

**Figure 2.**
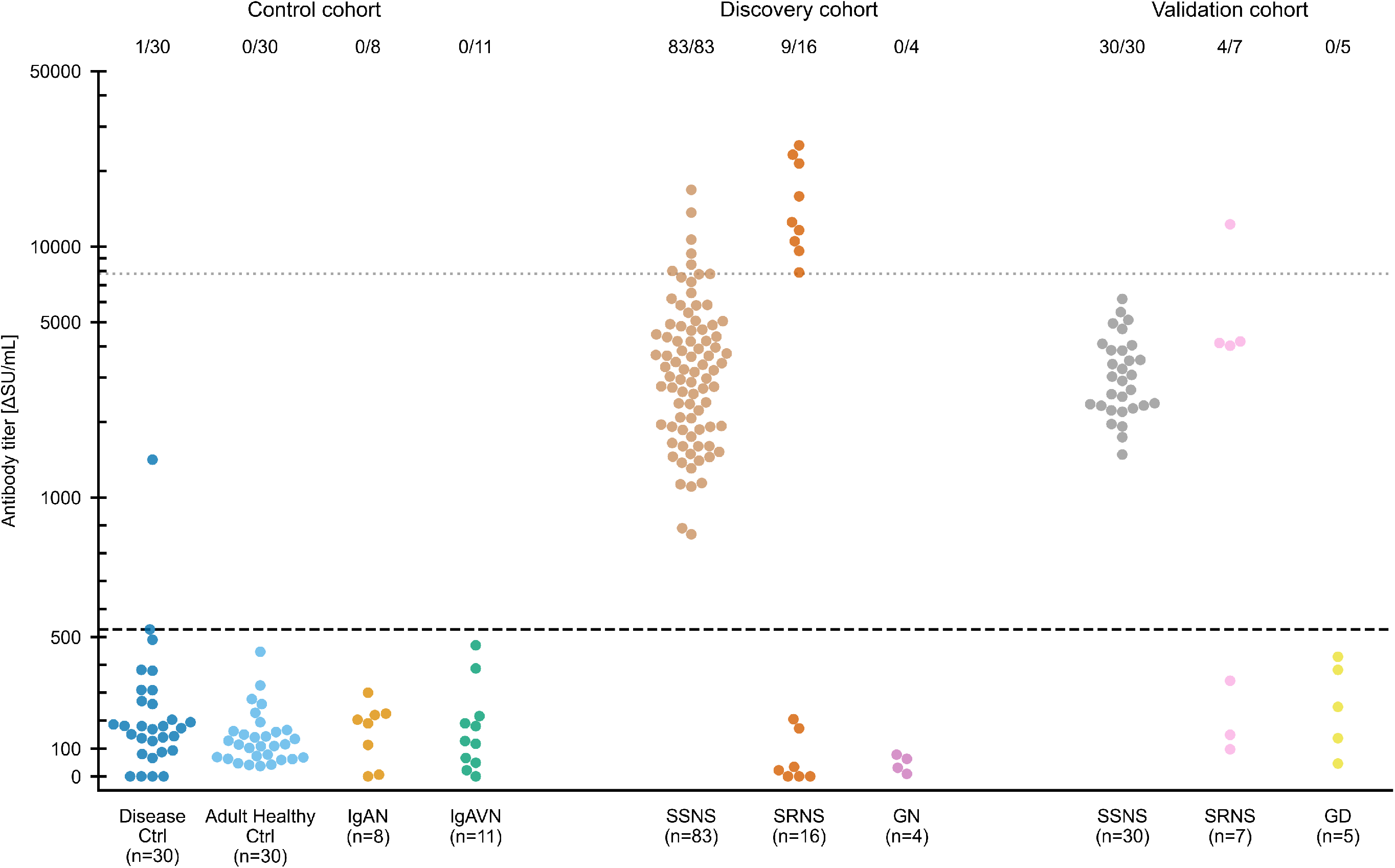
Distribution of pretreatment anti-nephrin autoantibody titers across pediatric nephrotic syndrome phenotypes and controls. Pretreatment anti-nephrin autoantibody titers are shown for the control, discovery, and validation cohorts. The control cohort included disease controls (n = 30), adult healthy controls (n = 30), biopsy-proven IgA nephropathy (IgAN, n = 8), and IgA vasculitis nephritis (IgAVN, n = 11). Patients in the discovery and validation cohorts were categorized according to their final diagnosis as steroid-sensitive nephrotic syndrome (SSNS), steroid-resistant nephrotic syndrome (SRNS), or diagnosed glomerulonephritis/other glomerular disease (GN/GD). Each dot represents an individual patient. Numbers above each group indicate the proportion of anti-nephrin autoantibody-positive cases. Horizontal dashed lines indicate the positivity cutoff (527 ΔSU/mL) and the high-titer cutoff (7,783 ΔSU/mL). The y-axis is plotted on a symmetrical logarithmic (symlog) scale, which is linear from 0 to 1,000 and logarithmic for values >1,000.

Table 1a summarizes anti-nephrin autoantibody titers measured before treatment initiation and the corresponding final diagnoses. Among the 103 patients, 92 (89.3%) tested positive for anti-nephrin autoantibodies. All patients with SSNS were antibody-positive, whereas all patients with GN were antibody-negative. In contrast, patients with SRNS demonstrated antibody-positivity (n = 9) and antibody-negativity (n = 7). Among the seven patients with SRNS who were negative for anti-nephrin autoantibodies, two patients failed to achieve remission with multiple immunosuppressive agents. These two patients underwent targeted next-generation sequencing, which revealed no pathogenic variants (Supplementary methods). In contrast, the remaining five patients with anti-nephrin autoantibody–negative SRNS ultimately achieved remission. All nine SRNS patients with positive anti-nephrin autoantibody titers achieved remission after immunosuppressive and/or steroid pulse therapy.

**Table 1.**
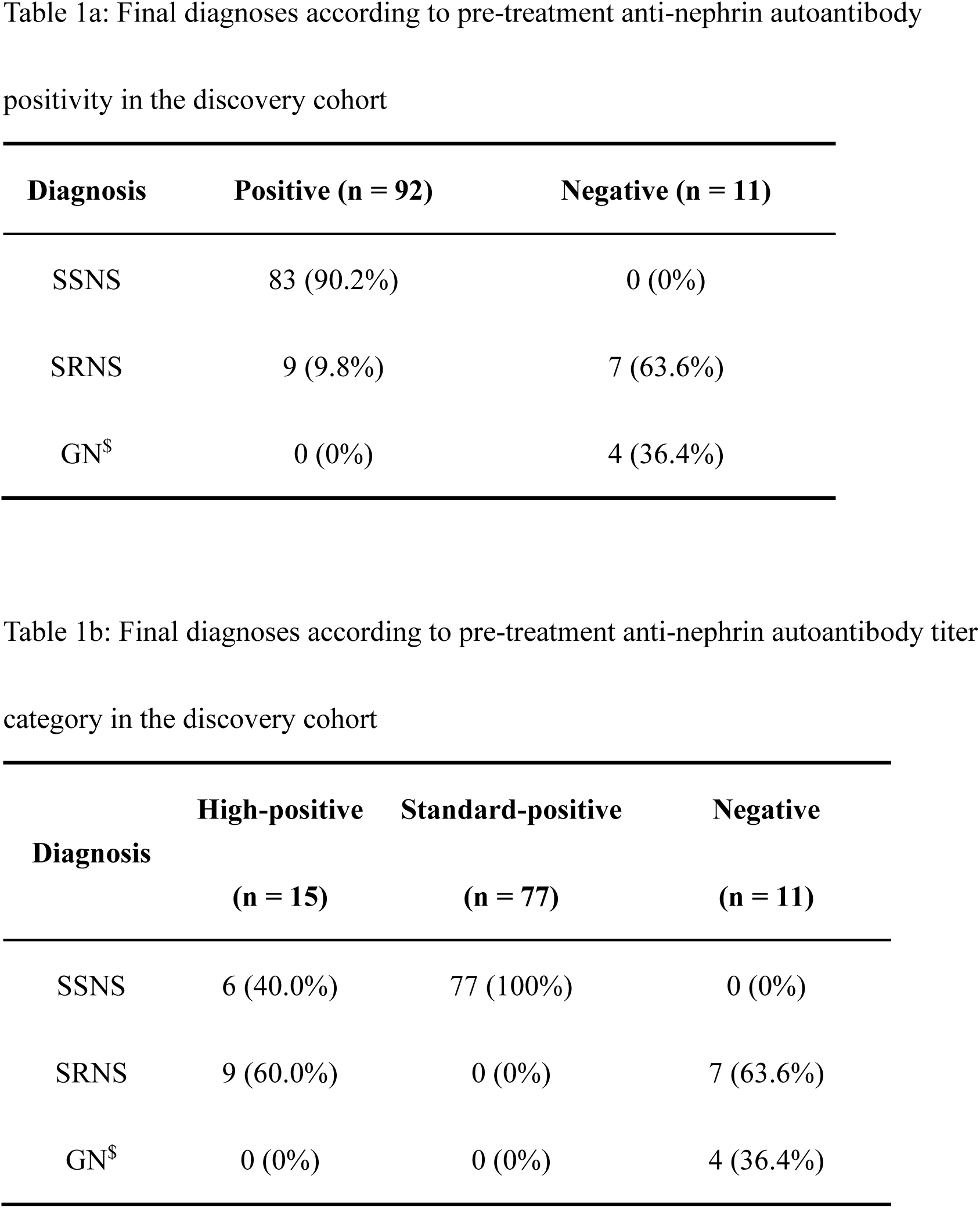
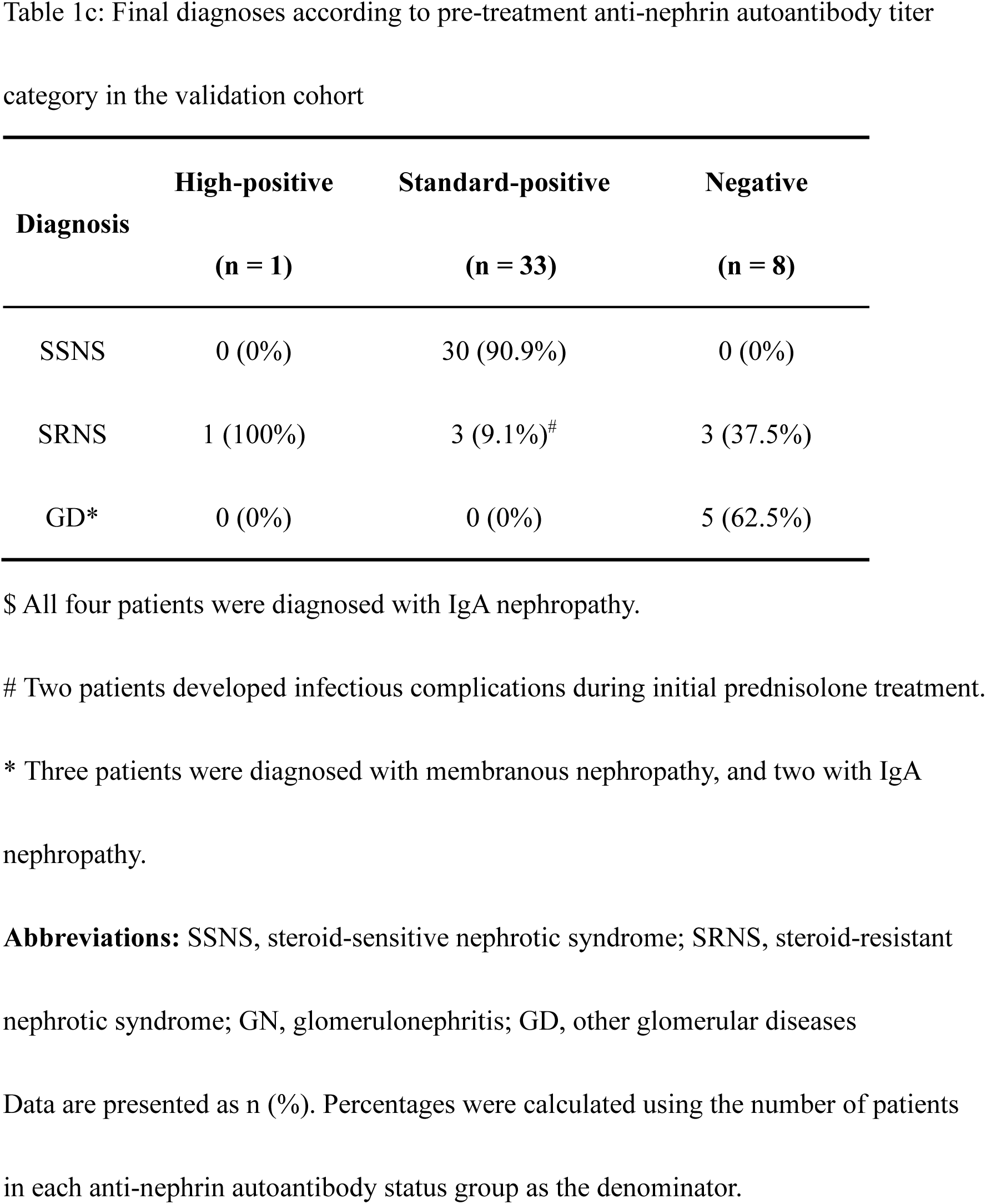
Final diagnoses according to pre-treatment anti-nephrin autoantibody status in the pediatric initial-onset nephrotic syndrome.

Among antibody-positive patients, the maximum Youden index for discriminating SSNS from SRNS was observed at a cut-off value of 7,783 ΔSU/mL, yielding a sensitivity of 100% and a specificity of 92.77% for identifying SSNS (Supplementary Methods and Supplementary Figure S6). Based on this cut-off value, patients were further classified into high-positive, standard-positive, and negative groups (Table 1b and Figure 2). All patients classified as standard-positive (n = 77) were ultimately diagnosed with SSNS. In contrast, among high-positive patients (n = 15), 9 were diagnosed with SRNS. Among antibody-negative patients (n = 11), 4 were diagnosed with IgAN, and the remaining 7 were diagnosed with SRNS. Standard-positive anti-nephrin autoantibody status was strongly associated with SSNS. In contrast, SRNS comprised patients with antibody-negativity and high-positivity, whereas GN was exclusively observed in antibody-negative patients. Thus, in children with newly diagnosed NS, anti-nephrin autoantibody status at disease onset was predictive of clinical outcome: positivity within the usual range was associated with SSNS, whereas both high-titer positivity and antibody negativity were associated with an increased risk of SRNS and other glomerular diseases (Fisher’s exact test, *P* < 0.0001).

Pre-treatment anti-nephrin autoantibody titers showed a weak positively significant correlation with time to remission in patients with SSNS (Spearman’s ρ = 0.264, 95% CI 0.045–0.460, *P* = 0.0160, n = 83), indicating that higher titers were associated with delayed remission (Figure 3).

**Figure 3.**
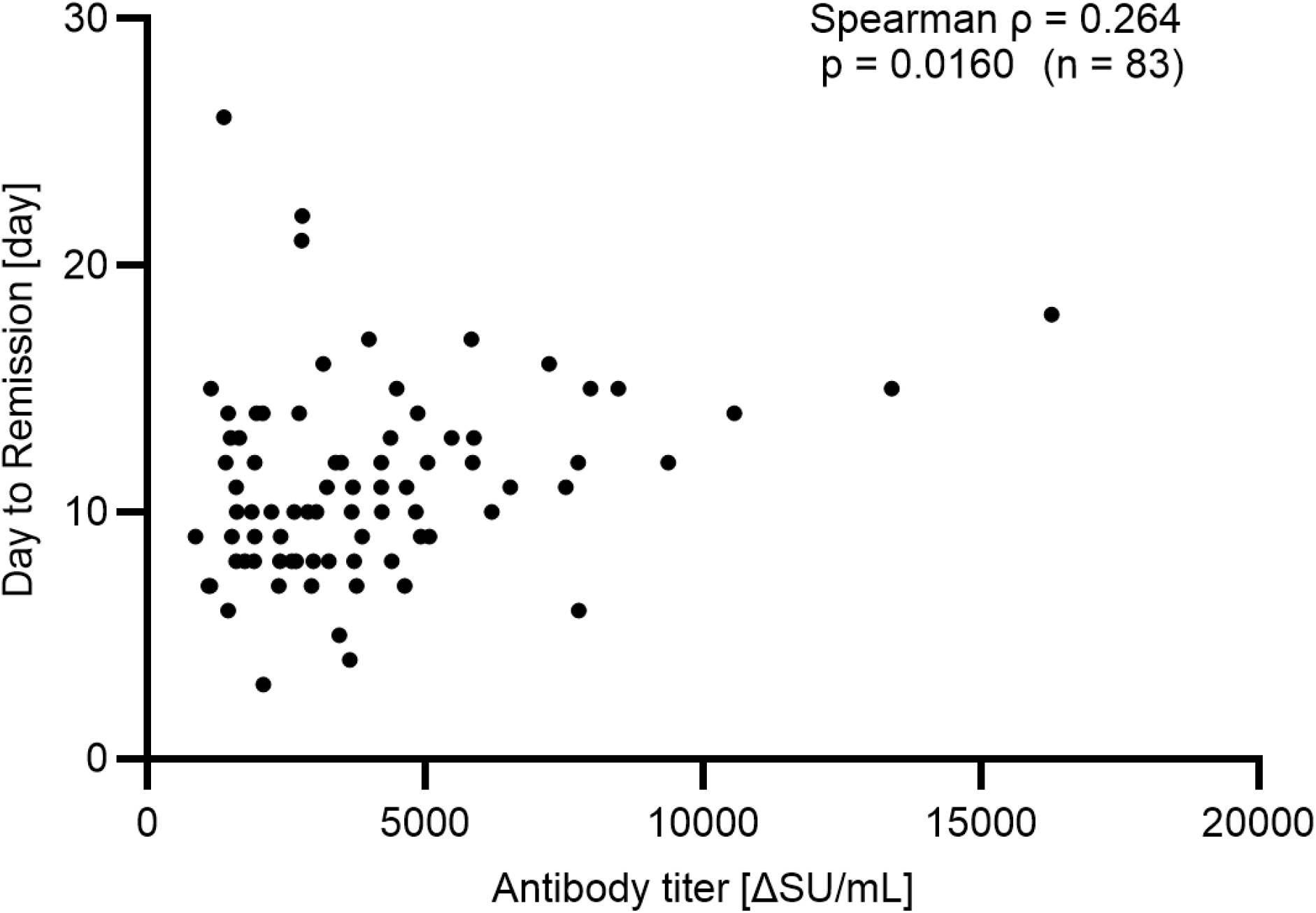
Correlation between pre-treatment anti-nephrin autoantibody titers and time to remission in steroid-sensitive nephrotic syndrome. Pre-treatment anti-nephrin autoantibody titers were positively correlated with time to remission in patients with steroid-sensitive nephrotic syndrome (SSNS) (Spearman’s ρ = 0.264, 95% CI 0.045–0.460, *P* = 0.0160, n = 83). Each point represents an individual patient. Higher antibody titers were associated with delayed remission.

#### Post-treatment anti-nephrin autoantibody titers in the pediatric discovery cohort

Supplementary Table S7. and Figure 4 show the anti-nephrin autoantibody status (positive/negative) in 91 patients with serum samples within 35 days after treatment initiation, and the slope graphs showing pre- and post-treatment changes in antibody titers for each patient. anti-nephrin autoantibody titers in patients with SSNS (n = 78) and SRNS (n = 11) decreased significantly after treatment (Wilcoxon matched-pairs signed-rank test, *P* < 0.0001 and *P* = 0.0020, respectively). Although a subset of patients with SSNS remained positive for anti-nephrin autoantibodies during remission, antibody-positive cases had a significantly shorter interval from the initial blood sampling to remission-phase sampling than antibody-negative cases (median 14 vs. 25 days; Mann–Whitney U test, *P* = 0.0069, Supplementary Figure S7).

**Figure 4.**
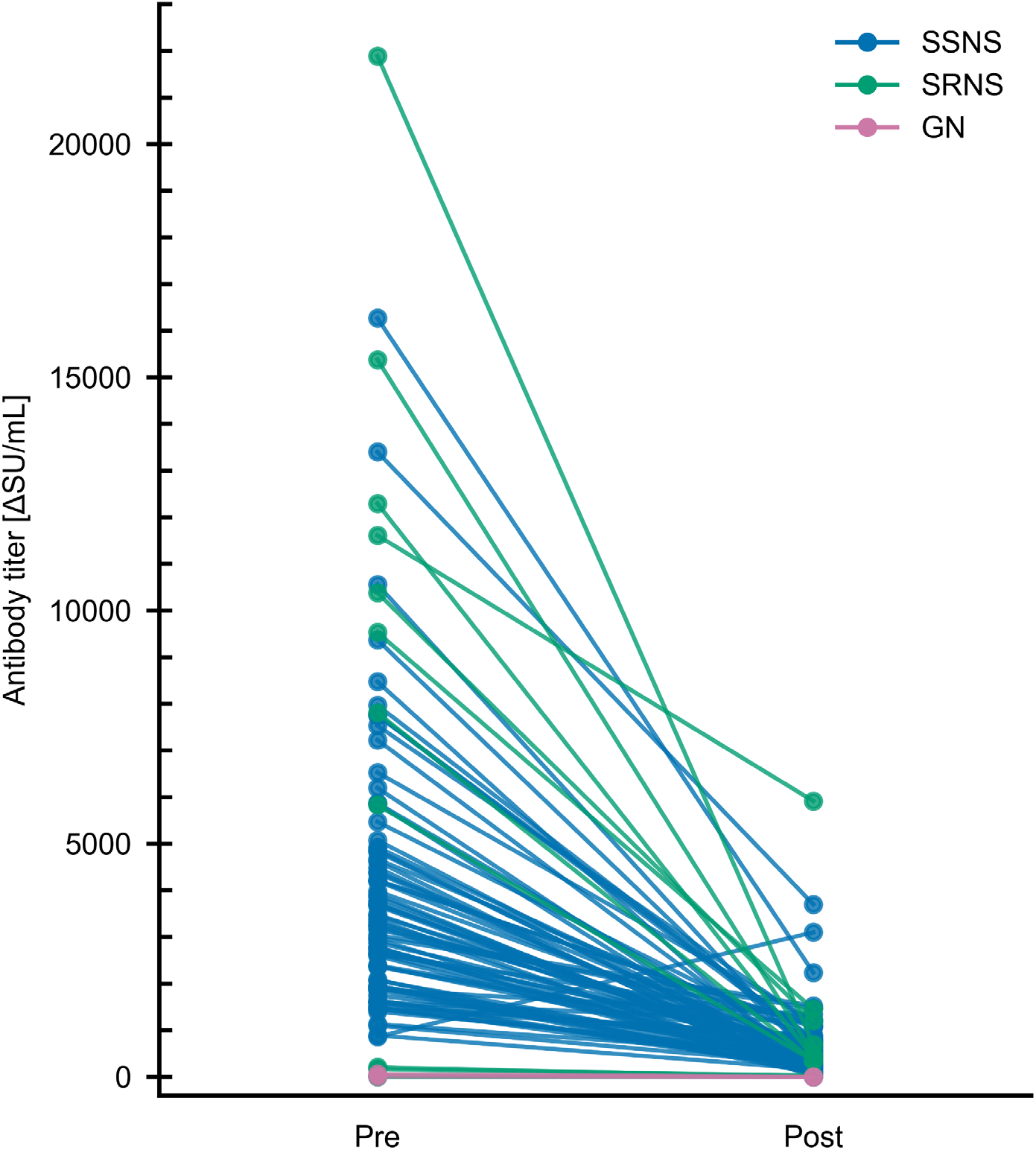
Changes in anti-nephrin autoantibody titers before and after treatment. Paired anti-nephrin autoantibody titers measured before treatment (Pre) and at remission or within 35 days after treatment initiation (Post) in patients with steroid-sensitive nephrotic syndrome (SSNS; blue, n = 78), steroid-resistant nephrotic syndrome (SRNS; green, n = 11), and glomerulonephritis (GN; red, n = 2). Each line represents an individual patient. Anti-nephrin autoantibody titers decreased significantly following treatment in both SSNS and SRNS patients (Wilcoxon matched-pairs signed-rank test, *P* < 0.0001 and *P* = 0.0020, respectively), whereas no significant change was observed in patients with GN.

#### Genetic risk alleles and anti-nephrin autoantibody response in the pediatric discovery cohort

Table 2 shows the distribution of *HLA* and *NPHS1* risk alleles across disease categories in the pediatric discovery cohort. *HLA* risk (rs3129888 G) alleles were more frequently observed in patients with SSNS than in those with SRNS or GN, although this difference was not statistically significant (Fisher’s exact test, *P* = 0.133). Similarly, the distribution of *NPHS1* risk (rs2285450 A) allele genotypes did not differ significantly among SSNS, SRNS, and GN (Fisher’s exact test, *P* = 0.186). The *NPHS1* risk allele was significantly more frequent in the SSNS group compared with controls ^8^ (OR = 1.93, 95% CI = 1.34–2.74; Fisher’s exact test; *P* < 0.001; Supplementary Table S8a).

**Table 2.** Distribution of *HLA* and *NPHS1* risk alleles according to final diagnosis in the pediatric initial-onset nephrotic syndrome discovery cohort.

|  | SSNS (n = 83) | SRNS (n = 16) | GN (n = 4) | *P*-value |
| --- | --- | --- | --- | --- |
| *HLA* risk allele carrier, n = 24 | 23 (27.7%) | 1 (6.3%) | 0 (0%) | 0.133 |
| *NPHS1* risk allele |  |  |  |  |
| Null, n = 63 | 49 (59.0%) | 13 (81.2%) | 1 (25.0%) | 0.186 |
| Heterozygous, n = 34 | 28 (33.7%) | 3 (18.8%) | 3 (75.0%) |  |
| Homozygous, n = 6 | 6 (7.2%) | 0 (0%) | 0 (0%) |  |

Abbreviations: GN, glomerulonephritis; SSNS, steroid-sensitive nephrotic syndrome; SRNS, steroid-resistant nephrotic syndrome.
Data are presented as n (%). Percentages were calculated within each diagnostic group.
*P*-values were calculated using Fisher's exact test.

Moreover, the *HLA* risk allele showed a marked enrichment in SSNS cases compared with controls ^32^ (OR = 4.57, 95% CI = 2.80–7.45; Fisher’s exact test; *P* < 0.0001) (Supplementary Table S8b). Among patients with SSNS, baseline anti-nephrin autoantibody titers did not differ between *HLA* risk allele carriers and non-carriers (Mann–Whitney U test, *P* = 0.38, Supplementary Figure S8a). Baseline anti-nephrin autoantibody titers did not differ among *NPHS1* risk allele genotype groups (null, heterozygous, and homozygous) with SSNS (Kruskal–Wallis test, *P* = 0.70; n = 83, Supplementary Figure S8b).

Supplementary Table S9. shows the genotype distributions in anti-nephrin autoantibody– positive and –negative groups. However, no significant association was observed between the groups.

#### Validation of pre-treatment anti-nephrin autoantibody findings in an independent pediatric cohort

We enrolled 42 children with newly diagnosed NS in the independent validation cohort. Among them, 1 patient was classified as high-positive, 33 as standard-positive, and 8 as negative for anti-nephrin autoantibodies at disease onset (Table 1c, Figure 2). All 30 patients who were ultimately diagnosed with SSNS were classified as standard positive. In contrast, among the 8 antibody-negative patients, 5 were diagnosed with alternative glomerular diseases (3 membranous nephropathy and 2 IgAN) and 3 with SRNS. The remaining 4 patients with SRNS were antibody-positive, including 1 high-positive and 3 standard-positive cases. Among the three patients with standard-positive SRNS, two developed infectious complications during treatment, which may have contributed to delayed remission induction. Notably, all patients with SSNS were classified as standard-positive, whereas all alternative glomerular diseases cases were antibody-negative, consistent with the findings in the discovery cohort.

#### Antigen inhibition assay in the validation cohort

We performed antigen inhibition assays in all antibody-positive samples from the validation cohort (SSNS, n = 30; SRNS, n = 4) and observed no marked signal changes in antibody-negative samples (disease control, n = 2; glomerular disease, n = 3; SRNS, n = 1; Adult healthy control, n = 4). In contrast, preincubation with recombinant nephrin antigen reduced antibody signals in all antibody-positive samples, including all SSNS cases and antibody-positive SRNS cases, supporting the specificity of the assay (Supplementary Figure S9).

## Discussion

In this study, we established a highly sensitive and automated immunoassay for the detection of anti-nephrin autoantibodies and prospectively evaluated anti-nephrin autoantibody titers in children with newly diagnosed NS. We found that pretreatment anti-nephrin autoantibody titers were strongly associated with clinical phenotype and treatment response. All patients with SSNS were antibody-positive, whereas non-SSNS patients, including SRNS and alternative glomerular diseases, most commonly exhibited either high-titer positivity or complete antibody negativity. These findings were reproduced in an independent validation cohort. Rather than representing a binary biomarker, anti-nephrin autoantibody titers could divide patients into distinct biological categories that differ in treatment responsiveness and disease phenotype.

Assay variability had been a major obstacle to the clinical application of anti-nephrin autoantibody testing.^17, 18^ By optimizing the recombinant nephrin antigen and implementing the assay on an automated chemiluminescent platform, we achieved highly sensitive and reproducible measurements with rapid turnaround that showed strong concordance with IP-WB analyses, kidney tissue immunostaining findings, and antigen inhibition assay results.

The clinical implications of these findings may extend beyond diagnosis. Some antibody-negative patients had other glomerular diseases, suggesting that a negative anti-nephrin autoantibody result may prompt consideration of alternative diagnoses and further diagnostic evaluation. Conversely, higher antibody titers were associated with longer remission times, indicating that anti-nephrin autoantibody burden may reflect disease activity. Serial monitoring may therefore provide additional information regarding treatment response and support future individualized treatment strategies. In contrast to previous reports,^12, 16^ all children with SSNS in our cohort were positive for anti-nephrin autoantibodies. Several factors may explain this exceptionally high detection rate. First, the optimized recombinant nephrin antigen and highly sensitive automated chemiluminescent platform may have improved the detection of low-level antibody responses that could have been missed by earlier assays. Second, all samples were obtained at initial presentation before corticosteroid and/or immunosuppressive therapy, thereby minimizing the potential effects of treatment on antibody detection as well as pathophysiological changes associated with prolonged disease duration. Although anti-nephrin autoantibody titers were not associated with the presence of *NPHS1* or *HLA* risk alleles, the relatively high frequency of nephrin-associated susceptibility alleles in the Japanese population^5, 8^ may have contributed to the remarkably high prevalence of anti-nephrin autoimmunity observed in this cohort. Antibody-negative SRNS patients appeared to constitute a biologically heterogeneous group. Patients who subsequently achieved remission following immunosuppressive therapy may represent alternative immune-mediated forms of podocytopathy, including those associated with autoantibodies against other podocyte proteins, such as podocin and KIRREL1,^13^ although these mechanisms were not evaluated in the present study, whereas those who remained treatment resistant may harbor underlying genetic or molecular abnormalities not detected by current diagnostic testing.

Some limitations should be acknowledged. First, the sample size was relatively small, particularly for patients with non-SSNS. Second, the study population consisted exclusively of Japanese patients, limiting the generalizability of the results. Further studies in ethnically diverse populations are warranted to determine the generalizability of these findings. Finally, although anti-nephrin autoantibodies were associated with clinical phenotypes, the present study was not designed to determine their direct pathogenic role.

In conclusion, our findings identify anti-nephrin autoimmunity as a central biological feature of SSNS and a subset of podocytopathies. Quantitative assessment of anti-nephrin autoantibody titers provides clinically relevant information that enables patient stratification and prognostic assessment. These data support a mechanism-based classification of NS and establish the automated immunoassay for anti-nephrin autoantibodies as a practical tool for future biomarker-guided management strategies. Further large-scale studies should be performed to validate this assay.

## Data Availability

The data that support the findings of this study are available from the corresponding author upon reasonable request.

## Acknowledgments

A patent application has been filed relating to this work. We are grateful to all patients and their families who participated in this study. We also acknowledge the invaluable contributions of all investigators and clinical research staff at the participating institutions. A full list of study investigators is provided in the Supplementary Materials.

## Author Contributions

TH, MM, AH, and KI conceived the study. TH, KN, KI, MM, MK, YTS, EMY, VP, JK, AT, YA, NN, KM, MO, AG, KK, and BDM developed the methodology. TH, CN, YIn, YIc, NS, TY, KF, KN, and KI conducted the investigation. TH and MM performed the formal analysis. TO, KKa, RH, NY, MG, HB, YK, RK, NF, HM, TK, RF, HK, EH, YIk, WS, YO, YS, TN, SA, MMi, KIsh, KNa, YI, and SM contributed to data curation. KI, KN, SM, and YI supervised the study. TH acquired funding. TH and MM wrote the original draft. AH reviewed and edited the manuscript. All authors reviewed and approved the final manuscript.

## Supplementary methods

### Plasmid construction

Genes encoding the WT human nephrin extracellular domain (CCC) and three triple-mutants with Cys to Val, Ala, or Ser substitutions at positions 101, 721, and 974 (VVV, AAA, and SSS, respectively) were synthesized and cloned into the pcDNA3.4 vector (Thermo Fisher Scientific, Waltham, MA). Based on the CCC sequence, single, double, and triple Val mutants (VCC, CVC, CCV, CVV, VCV, VVC, and VVV) were generated via PCR-based site-directed mutagenesis using synthesized primers. The resulting PCR products were fragmented with DpnI (Toyobo, Osaka, Japan, DPN-101), phosphorylated using T4 Polynucleotide Kinase (Takara Bio, Shiga, Japan, 2021A), and ligated using Ligation High-Ver.2 (Toyobo, LGK-201).

The generated plasmids and ligation mixtures were transformed into Competent high DH5α cells (Toyobo, DNA-913F). Single colonies were cultured in liquid media, and plasmids were extracted using a Maxiprep Kit (QIAGEN, Hilden, Germany, 12123). All nucleotide sequences were verified by Sanger sequencing using a pcDNA3.4 vector primer.

### Protein expression and purification

Expi293F cells (Thermo Fisher Scientific, A14527) were maintained in Expi293 Expression Medium (Thermo Fisher Scientific, A1435101) at 37 °C and 5% CO2 with shaking at 125 rpm. Cells were transfected with the prepared plasmids using the ExpiFectamine 293 Transfection Kit (Thermo Fisher Scientific, A14524). Twenty hours post-transfection, we added ExpiFectamine Transfection Enhancers 1 and 2 to the culture, and the cells were cultured for an additional 4 days.

The cell culture was centrifuged at 3,000 rpm for 30 minutes to collect the supernatant, which was subsequently frozen in liquid nitrogen and stored at −80 °C. For purification, the supernatant was thawed under running water and dialyzed against a phosphate buffer containing 40 mM imidazole using a Slide-A-Lyzer Dialysis Cassette (Thermo Fisher Scientific, A52980). The dialyzed sample was loaded onto a HisTrap HP column (Cytiva, Marlborough, MA, 17524701) connected to an AKTA pure system (Cytiva). The column was washed with the 40 mM imidazole phosphate buffer, and the recombinant proteins were eluted using a phosphate buffer containing 300 mM imidazole. The eluted proteins were stored until use in subsequent experiments.

### Establishment of an automated immunoassay for anti-nephrin autoantibody detection

The chemiluminescence enzyme immunoassay was performed using a 4-(2-hydroxyethyl)-1-piperazineethanesulfonic acid (HEPES)-based sample diluent and the following five (HISCL reagents (R1–R5):

- **R1 reagent:** HEPES buffer (pH 6.5).
- **R2 reagent:** Magnetic particles coated with either wild-type (CCC) or mutated human nephrin extracellular domains in which three free cysteine residues were substituted (SSS, AAA, VVV) in HEPES buffer (pH 6.5).
- **R3 reagent:** In-house alkaline phosphatase (ALP)-fused mouse anti-human IgG monoclonal antibody in HEPES buffer (pH 6.5).
- **R4 reagent:** Assay buffer (HISCL R4; Sysmex, Kobe, Japan).
- **R5 reagent:** Chemiluminescent substrate solution containing CDP-Star (HISCL R5; Sysmex).

### Optimization of the nephrin antigen

To optimize the antigen for the assay, we conducted a two-step evaluation. First, to determine the optimal substituting amino acid, we compared the immunoreactivities of CCC and the three triple-mutants in which free cysteine residues were substituted (VVV, AAA, and SSS) using clinical samples from patients with pediatric nephrotic syndrome. Subsequently, to identify the optimal substitution positions, we evaluated CCC alongside the single and double Val mutants, as well as VVV, using samples from pediatric nephrotic syndrome patients and healthy controls. Based on these evaluations, the triple Val-substituted mutant was selected for the final assay formulation.

### Western blotting analysis of recombinant nephrin variants under reducing and non-reducing conditions

Purified samples were mixed with reducing SDS-PAGE sample buffer (Nacalai Tesque, Kyoto, Japan, 09499-14) or non-reducing SDS-PAGE sample buffer (Nacalai Tesque, 09500-64), followed by heating at 96 °C for 5 min. The samples were separated by SDS-PAGE and transferred to polyvinylidene difluoride membranes using the iBlot 2 Dry Blotting System and iBlot 2 Transfer Stacks, PVDF (Thermo Fisher Scientific, IB24001). After transfer, the membranes were blocked with Block Ace Powder (KAC, Kyoto, Japan,UKB80), prepared according to the manufacturer’s instructions, for 1.5 h at room temperature. The membranes were then incubated overnight at 4 °C with a polyclonal sheep anti-nephrin antibody recognizing both human and mouse nephrin (R&D Systems, Minneapolis, MN, AF4269), diluted 1:500 in tenfold-diluted Block Ace. After three washes with Tris-buffered saline containing 0.1% Tween 20 (TBS-T), the membranes were incubated for 1 h at room temperature with horseradish peroxidase-conjugated donkey anti-sheep IgG (Jackson ImmunoResearch, West Grove, PA, AB_234071), diluted 1:100,000 in tenfold-diluted Block Ace. The membranes were washed three additional times with TBS-T.

Chemiluminescent signals were developed using SuperSignal West Pico PLUS substrate (Thermo Fisher Scientific, 34585), and images were acquired at multiple exposure settings using a FUSION SOLO S imaging system (Vilber GmbH, Collégien, France).

### Preparation of antigen-coated magnetic particles (R2 reagent)

Recombinant human nephrin antigens (CCC, and free cysteine-substituted variants SSS, AAA, or VVV) were immobilized onto carboxylated magnetic particles (3 μm in diameter) via the amine coupling method. The carboxyl groups on the particle surface were activated to form N-hydroxysuccinimide esters, followed by the addition of the recombinant nephrin proteins to form covalent bonds. The prepared R2 reagents were flash-frozen in liquid nitrogen and stored at −80°C until use. The reagents were thawed under running water immediately before loading. BSA-coated magnetic particles were prepared in the same manner by immobilizing BSA instead of the recombinant nephrin antigens to calculate the ΔSU/mL.

### Measurement protocol

The measurement was conducted according to the following manual and HISCL-5000 system-based steps:

1. **Manual sample pre-processing:** Serum or plasma samples were diluted 40-fold with a HEPES-based sample diluent (pH 6.5) and then filtered through a 0.22-μm filter.
2. **Primary reaction (Step 1):** The filtered sample was first mixed with the R1 reagent. This mixture was then reacted with the R2 reagent (antigen-coated magnetic particles) to capture anti-nephrin autoantibodies.
3. **Washing:** After magnetic separation, the particles were washed with HISCL Wash Solution (Sysmex) to remove unbound IgG and other matrix components.
4. **Secondary reaction (Step 2):** The R3 reagent (ALP-fused anti-human IgG; Sysmex) was added to the particles to detect the captured autoantibodies.
5. **Final washing and detection:** After a second washing step, the R4 and R5 reagents were added and mixed.
6. **Signal measurement:** The chemiluminescence intensity was measured, and the antibody titer was calculated based on the luminescence signal.

### Calibration curve

An assay-specific unit, designated as SU/mL, was adopted for quantitation since the measurement target is an autoantibody and the signal depends on its binding affinity. The calibrators were prepared by serially diluting a high-titer sample by weight using a HEPES-based sample diluent. The calibration curve consisted of five or six points (a blank and four or five concentration points) and was constructed using a four-parameter logistic regression model to convert the chemiluminescence intensity (HISCL counts) into SU/mL.

### Determination of the quantitative range

To establish the quantitative range of the assay, the limit of quantitation (LoQ; the lower limit) and the upper limit of quantitation (ULoQ) were determined. A sample from a pediatric patient with nephrotic syndrome was serially diluted with the HEPES-based sample diluent to define the LoQ. Each dilution point was measured in five replicates (n = 5). The LoQ was defined as the lowest concentration at which the coefficient of variation 15% or less. To determine the ULoQ, dilution linearity was evaluated using three high-titer samples from pediatric patients with nephrotic syndrome. These samples were serially diluted with the HEPES-based sample diluent, and each dilution point was measured in two or three replicates (n = 2–3). The ULoQ was defined as the highest concentration that maintained dilution linearity.

### Intra- and inter-assay reproducibility

To evaluate the precision of the assay, intra-assay and inter-assay reproducibility were assessed using three serum samples from pediatric patients with nephrotic syndrome. These samples represented low, middle, and high anti-nephrin autoantibody concentrations. For the assessment of intra-assay reproducibility, each of the three samples was measured in 10 replicates (n = 10) within a single assay run. To evaluate inter-assay reproducibility, each sample was measured once a day over three days (n = 3).

### Sample stability

To evaluate sample stability, two samples from pediatric patients with nephrotic syndrome (one with a low and one with a high antibody concentration) were aliquoted. To assess the effects of storage conditions, the aliquots were stored at −80°C (as a control), −20°C for 7 days, 4°C for 1 day, or 25°C for 1 day. Furthermore, to evaluate the impact of repeated freezing and thawing, aliquots were subjected to 1, 3, and 5 freeze-thaw cycles. A single freeze-thaw cycle was defined as maintaining the sample at room temperature for at least 30 minutes, followed by freezing at −80°C for at least 3 hours. All samples were measured in triplicate (n = 3).

### Validation of sample types

Paired serum and plasma samples from 20 pediatric patients with nephrotic syndrome (14 in the active phase and 6 in remission) were analyzed to verify the comparability of measurement values between sample types. These measurements were conducted using the optimized protocol, which incorporated the 0.22-μm filtration step following sample dilution. All samples were measured in duplicate (n = 2).

### Interference substances

“Interference Check A Plus” and “Interference Check RF Plus” (Sysmex) were used to evaluate the influence of interfering substances on the assay values. For bilirubin F (free), bilirubin C (conjugated), hemolytic hemoglobin, and chyle, each interfering substance was reconstituted with purified water and then mixed with serum samples from pediatric patients with nephrotic syndrome at a ratio of 9:1 to prepare spiked samples. As a control, the same sample was mixed with a blank solution at a ratio of 9:1. These mixtures were diluted 40-fold with the sample diluent and filtered through a 0.22-μm membrane before measurement.

The rheumatoid factor (RF) reagent or blank was reconstituted with the sample diluent, and a 39-fold diluted solution was prepared using the same diluent. Using this solution, serum samples from pediatric patients with nephrotic syndrome were diluted 40-fold and filtered through a 0.22-μm membrane.

In all evaluations, only the maximum concentration specified by the kits for each interfering substance was tested, and spiked samples (or RF-spiked samples) and control samples (blanks) were each measured in five replicates (n = 5). The maximum concentrations evaluated (calculated as concentrations in the original specimen) were as follows:

- **Bilirubin F**: 20.1 mg/dL
- **Bilirubin C**: 19.7 mg/dL
- **Hemolytic hemoglobin**: 510 mg/dL
- **Chyle**: 1,430 FTU
- **Rheumatoid factor (IgM-RF)**: 450 IU/mL

### Detection of anti-nephrin autoantibodies by immunoprecipitation–Western blotting (IP-WB)

Anti-nephrin antibodies were evaluated by IP-WB based on previously reported methods^1, 2^ with minor modifications. Briefly, 30 μL of patient serum was incubated with 150 ng of recombinant human nephrin protein in RIPA buffer (150 mM NaCl, 1% NP-40, 0.5% sodium deoxycholate, 0.1% SDS, and 50 mM Tris-HCl, pH 7.4) at 4°C overnight with rotation.

Following incubation, Protein G agarose resin (GenScript, Piscataway, NJ, L00209) was added and further incubated at 4°C for 1.5 h with rotation to capture patient IgG-bound nephrin complexes. The resin was subsequently washed twice with RIPA buffer and three times with TBS-T.

Bound proteins were eluted under reducing conditions by adding SDS-PAGE sample buffer containing SDS, reducing agent, and 100 mM dithiothreitol (DTT), followed by heating at 95°C for 10 min. Eluted proteins were separated by SDS-PAGE and transferred onto PVDF membranes using the iBlot 2 Gel Transfer Device and iBlot 2 Transfer Stacks, PVDF (Thermo Fisher Scientific, IB24001).

Membranes were blocked with Block Ace (KAC Co., UKB80) dissolved according to the manufacturer’s instructions for 1.5 h at room temperature and then incubated overnight at 4°C with a polyclonal sheep anti-nephrin antibody recognizing both human and mouse nephrin (R&D Systems, AF4269) diluted 1:200 in blocking buffer diluted tenfold with water. After three washes with TBS-T, membranes were incubated for 1 h at room temperature with HRP-conjugated donkey anti-sheep IgG antibody (Jackson ImmunoResearch, AB_234071) diluted 1:20,000 in blocking buffer diluted tenfold with water. Membranes were washed three additional times with TBS-T.

Chemiluminescent signals were detected using SuperSignal West Pico Plus substrate (Thermo Fisher Scientific, 34585) and visualized using a FUSION SOLO S imaging system (Vilber GmbH, Collégien, France).

### Targeted panel sequence

Targeted sequencing using next-generation sequencing was conducted for genes responsible for inherited kidney diseases, which included genes causing SRNS as previously described.^3^ The sample library for next-generation sequencing was prepared using the HaloPlex or SureSelect XT2 custom capture library 0.5–2.9 Mb (Agilent Technologies, Santa Clara, CA, USA), in accordance with the manufacturer’s workflow. All indexed DNA samples were amplified by polymerase chain reaction and sequenced using the MiSeq platform (Illumina, San Diego, CA, USA). Variant calling was performed according to the Genome Analysis Toolkit Best Practices workflows developed by the Broad Institute (Cambridge, MA),^4^ and variants were annotated using ANNOVAR.^5^

### Determination of the optimal anti-nephrin autoantibody cut-off by ROC analysis for distinguishing SSNS from SRNS

Receiver operating characteristic (ROC) curve analysis was performed to determine the cutoff value for predicting the risk of developing SRNS. The optimal cut-off value was defined as the point maximizing the Youden index (sensitivity+specificity−1).

---

**Supplementary Table S1.**
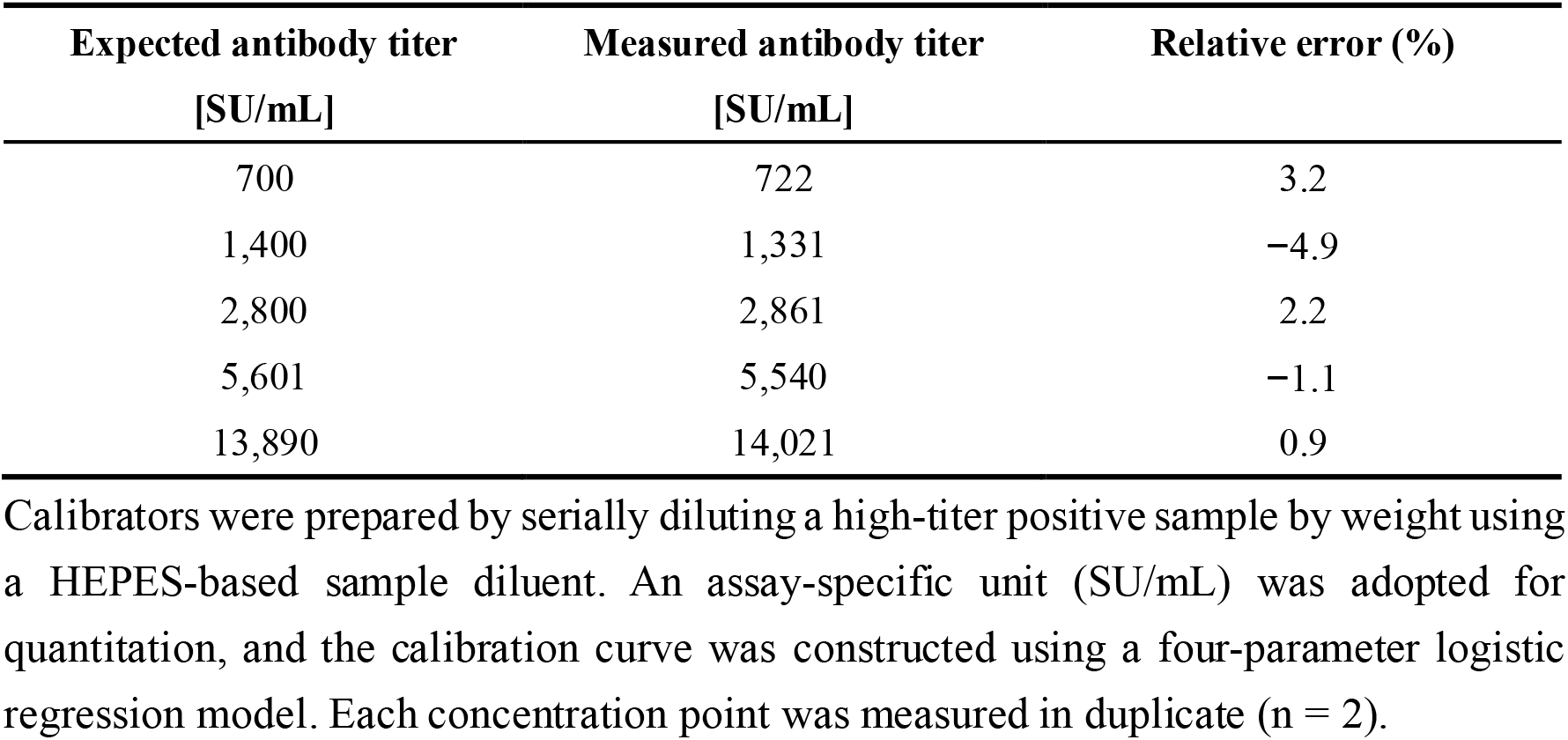
Accuracy of the calibration curve.

**Supplementary Table S2.** Determination of the limit of quantitation (LoQ)

| Dilution | Measured values [SU/mL] |  |  |  |  | Mean [SU/mL] | CV (%) |
| --- | --- | --- | --- | --- | --- | --- | --- |
|  | 1 | 2 | 3 | 4 | 5 |  |  |
| C1 | 696.3 | 673.9 | 677.0 | 678.1 | 696.0 | 684.3 | 1.6 |
| 1/2 C1 | 323.1 | 351.7 | 337.4 | 350.8 | 337.2 | 340.0 | 3.5 |
| 1/4 C1 | 143.1 | 155.9 | 159.3 | 156.2 | 145.3 | **152.0** | 4.8 |
| 1/8 C1 | 59.5 | 74.0 | 80.7 | 56.5 | 73.9 | 68.9 | **15.1** |

A sample from a pediatric patient with nephrotic syndrome was serially diluted with a HEPES-based sample diluent to define the LoQ. Each dilution point was measured in five replicates ( $n = 5$ ). The LoQ was defined as the lowest concentration at which the coefficient of variation (CV) was 15% or less. Based on this criterion, the LoQ was determined to be 152.0 SU/mL.

**Supplementary Table S3.** Intra- and inter-assay reproducibility of the anti-nephrin antibody assay.

| Sample | Antibody titer<br>[SU/mL] | Intra-assay<br>CV [%] | Inter-assay<br>CV [%] |
| --- | --- | --- | --- |
| Low | 1423 | 1.7 | 3.9 |
| Middle | 4480 | 1.1 | 5.4 |
| High | 6930 | 1.7 | 3.1 |

Three serum samples from pediatric patients with nephrotic syndrome, representing low, middle, and high anti-nephrin autoantibody concentrations, were used to evaluate assay precision. For intra-assay reproducibility, each sample was measured in 10 replicates within a single assay run ( $n = 10$ ). For inter-assay reproducibility, each sample was measured once a day over three days ( $n = 3$ ). The antibody titer presented in the table represents the mean concentration derived from the inter-assay measurements.

**Supplementary Table S4.** Influence of interfering substances on the anti-nephrin autoantibody assay.

| Interfering substances | Concentrations | Bias[%] |
| --- | --- | --- |
| Hemolytic hemoglobin | 510 mg/dL | -4.5 |
| Chyle | 1430 FTU | -6.4 |
| Free bilirubin | 20.1 mg/dL | -1.2 |
| Conjugated bilirubin | 19.7 mg/dL | 3.1 |
| Rheumatoid factor | 450 IU/mL | -2.7 |

The interference of major blood components on the assay values was evaluated at their maximum concentrations. Spiked samples were prepared by mixing each interfering substance with serum samples from pediatric patients with nephrotic syndrome. Control samples were mixed with a blank solution. All evaluations were measured in five replicates (n = 5). Bias represents the percentage deviation of the spiked samples compared to the control samples.

**Supplementary Table S5.**
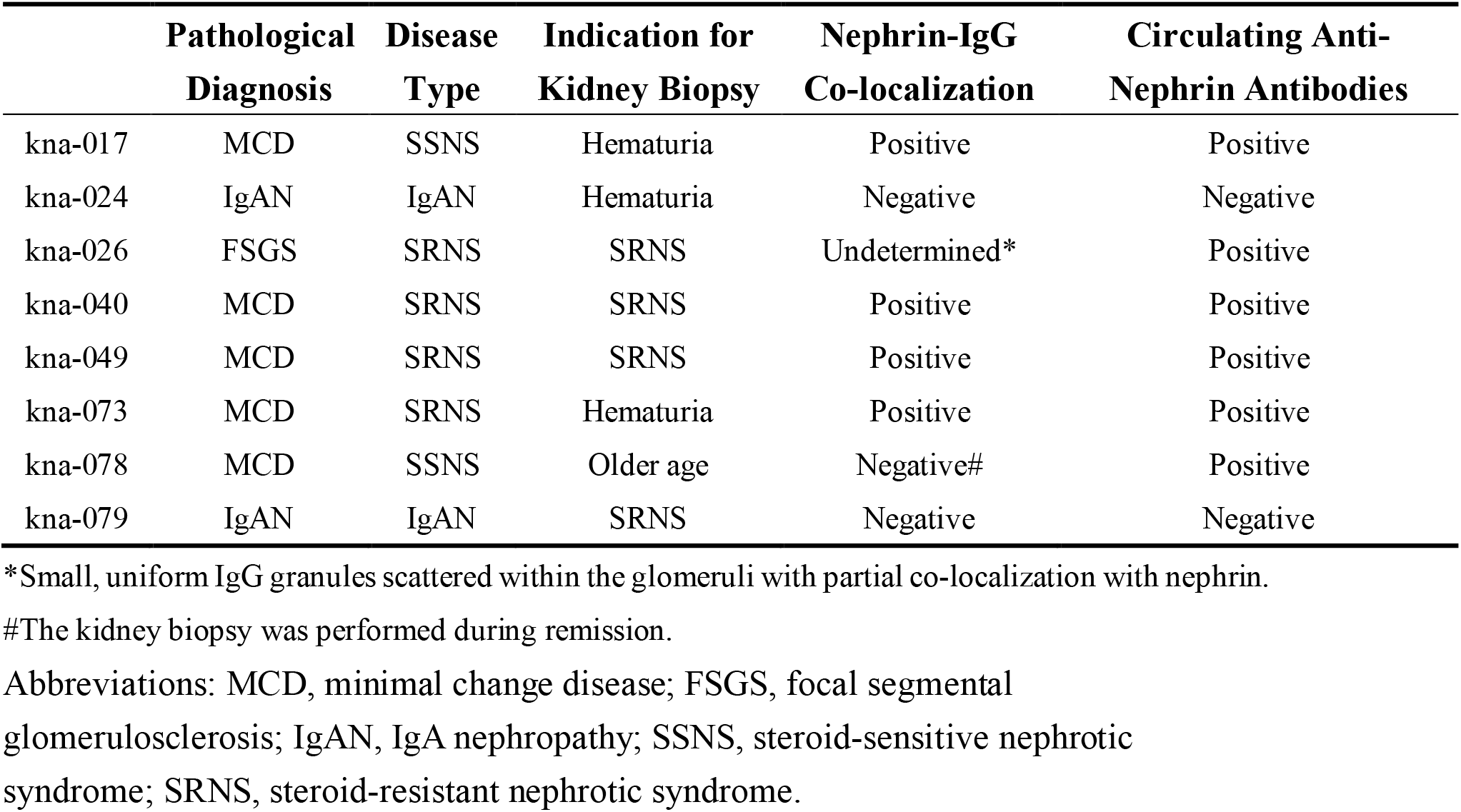
Clinicopathological findings and anti-nephrin autoantibody status in patients who underwent kidney biopsy.

|  | **Pathological**<br>**Diagnosis** | **Disease**<br>**Type** | **Indication for**<br>**Kidney Biopsy** | **Nephrin-IgG**<br>**Co-localization** | **Circulating Anti-**<br>**Nephrin Antibodies** |
| --- | --- | --- | --- | --- | --- |
| kna-017 | MCD | SSNS | Hematuria | Positive | Positive |
| kna-024 | IgAN | IgAN | Hematuria | Negative | Negative |
| kna-026 | FSGS | SRNS | SRNS | Undetermined* | Positive |
| kna-040 | MCD | SRNS | SRNS | Positive | Positive |
| kna-049 | MCD | SRNS | SRNS | Positive | Positive |
| kna-073 | MCD | SRNS | Hematuria | Positive | Positive |
| kna-078 | MCD | SSNS | Older age | Negative# | Positive |
| kna-079 | IgAN | IgAN | SRNS | Negative | Negative |

\*Small, uniform IgG granules scattered within the glomeruli with partial co-localization with nephrin.
#The kidney biopsy was performed during remission.
Abbreviations: MCD, minimal change disease; FSGS, focal segmental glomerulosclerosis; IgAN, IgA nephropathy; SSNS, steroid-sensitive nephrotic syndrome; SRNS, steroid-resistant nephrotic syndrome.

**Supplementary Table S6.** Baseline clinical characteristics of pediatric patients in the discovery cohort.

|  | Unit | Value |
| --- | --- | --- |
| Sex (Male/Female) | n | 73/30 |
| Age | months | 66 (34–120) |
| WBC | /μL | 8,900 (6,750–12,400) |
| Lymphocytes | /μL | 3,321 (2,574–4,754) |
| Hemoglobin | g/dL | 13.4 (12.6–14.6) |
| Platelets | $\times 10^4$/μL | 38.9 (31.3–47.9) |
| Total protein | g/dL | 4.0 (3.6–4.6) |
| Albumin | g/dL | 1.2 (0.9–1.8) |
| IgG | mg/dL | 237 (140–423) |
| Blood urea nitrogen | mg/dL | 12.1 (9.4–17.0) |
| Creatinine | mg/dL | 0.33 (0.25–0.53) |
| Urinary protein-to-creatinine ratio | g/gCr | 12.2 (7.3–19.0) |
| Hematuria, n (%) |  | 41 (39.8) |

Abbreviations: BUN, blood urea nitrogen; IgG, immunoglobulin G; WBC, white blood cell count; U-TP/Cr, urinary protein-to-creatinine ratio.
Data are presented as median (interquartile range) or n (%). Hematuria was defined as the presence of urinary red blood cells on routine urinalysis.

**Supplementary Table S7.** Anti-nephrin autoantibody status in post-treatment samples according to final diagnosis in the pediatric initial-onset nephrotic syndrome discovery cohort.

| Diagnosis | Positive (n = 22) | Negative (n = 69) |
| --- | --- | --- |
| SSNS | 19 (24.4%) | 59 (75.6%) |
| SRNS | 3 (27.3%) | 8 (72.7%) |
| GN | 0 (0%) | 2 (100%) |

Abbreviations: GN, glomerulonephritis; SSNS, steroid-sensitive nephrotic syndrome; SRNS, steroid-resistant nephrotic syndrome.
Data are presented as n (%). Percentages were calculated using the number of patients in each diagnostic category as the denominator. Post-treatment samples were obtained at remission or within 35 days after treatment initiation.

**Supplementary Table S8.**
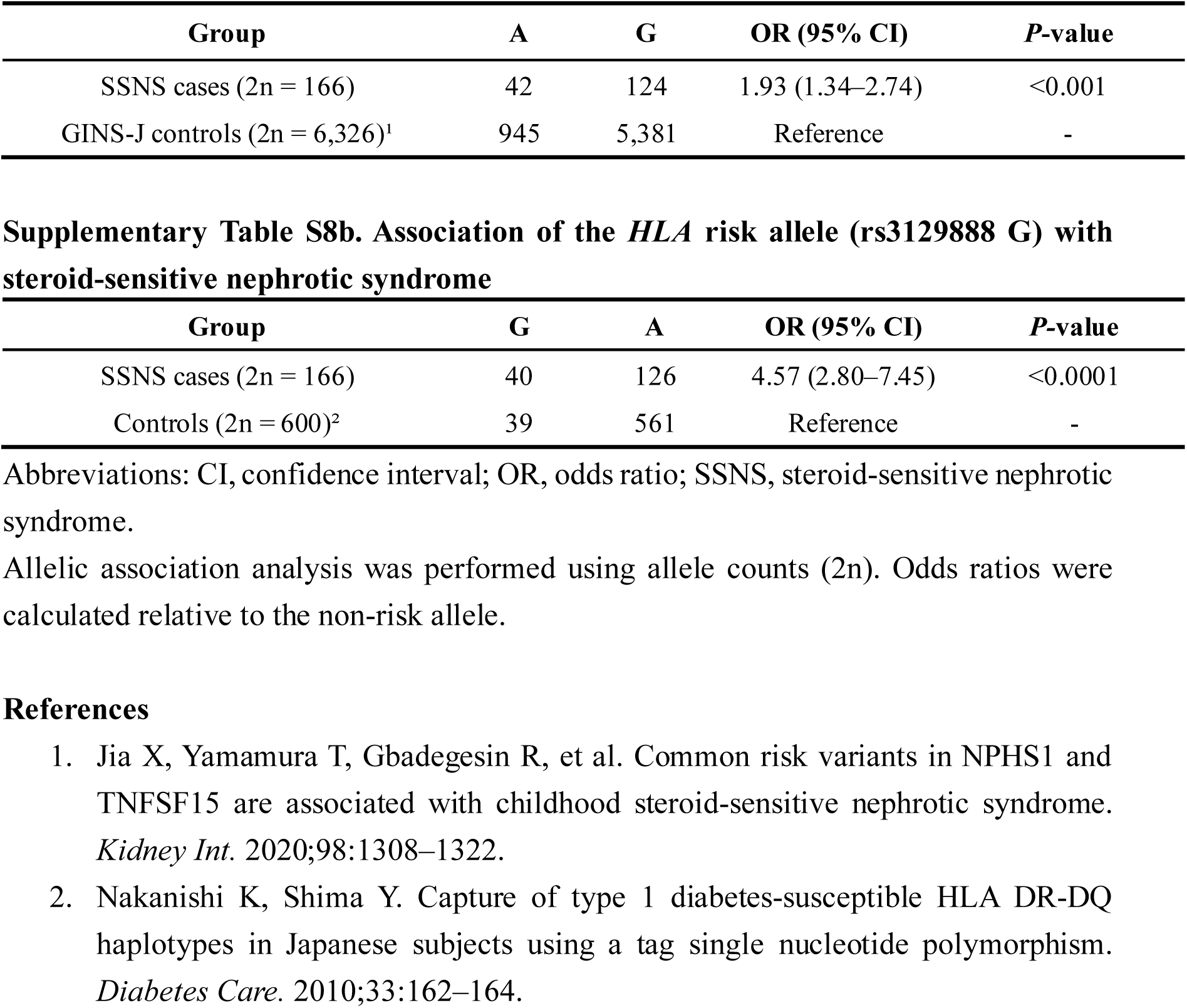
a. Association of the *NPHS1* risk allele (rs2285450 A) with steroid-sensitive nephrotic syndrome.

**Supplementary Table S9.** Distribution of *HLA* and *NPHS1* risk alleles according to anti-nephrin autoantibody status in the pediatric initial-onset nephrotic syndrome discovery cohort.

|  | Positive (n = 92) | Negative (n = 11) | *P*-value |
| --- | --- | --- | --- |
| *HLA* risk allele carrier, n = 24 | 23 (25.0%) | 1 (9.1%) | 0.45 |
| *NPHS1* risk allele |  |  |  |
| Null, n = 63 | 56 (60.9%) | 7 (63.6%) | 0.68 |
| Heterozygous, n = 34 | 30 (32.6%) | 4 (36.4%) |  |
| Homozygous, n = 6 | 6 (6.5%) | 0 (0%) |  |

Data are presented as n (%). Percentages were calculated using the number of patients in each anti-nephrin autoantibody status group as the denominator. *P*-values for *HLA* risk allele carrier status and *NPHS1* genotype distribution were calculated using Fisher's exact test.

**Supplementary Figure S1.**
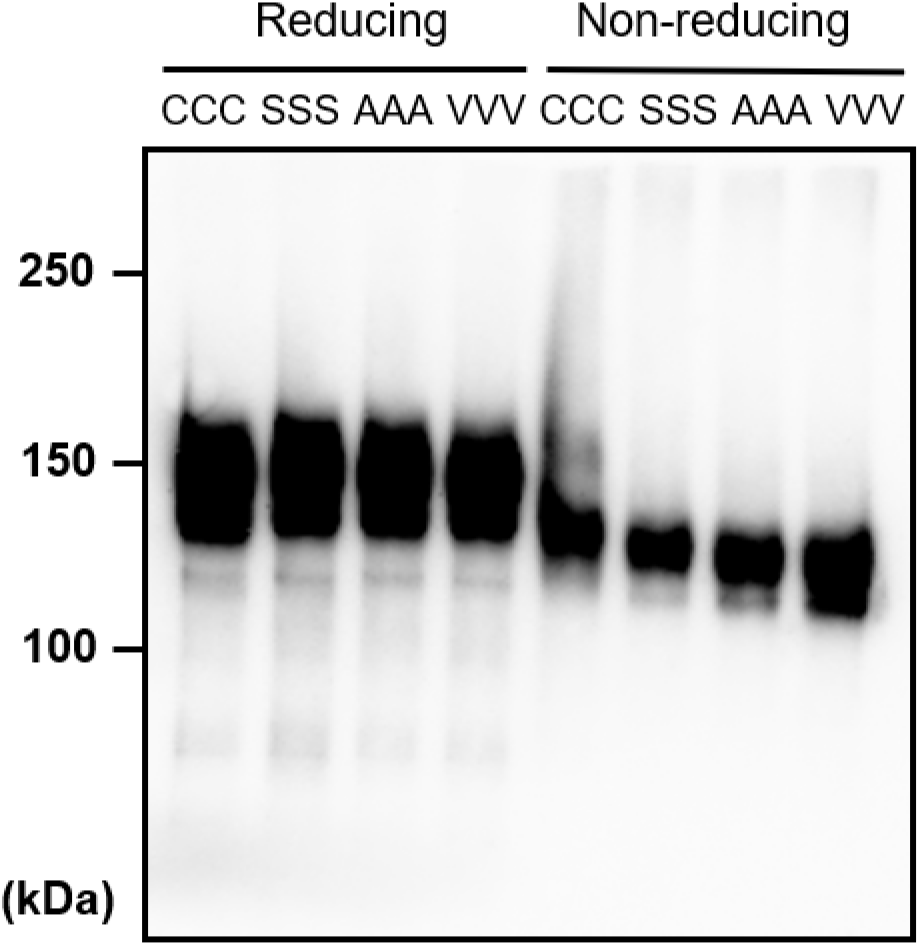
Electrophoretic migration of recombinant nephrin variants under reducing and non-reducing conditions. Immunoblot with a polyclonal anti-nephrin antibody showing the relative molecular migration of nephrin purified from transiently transfected Expi293F cells expressing the WT human nephrin extracellular domain (CCC) or one of three mutant variants in which all three free cysteine residues were substituted with serine (SSS), alanine (AAA), or valine (VVV), respectively, under reducing and non-reducing conditions.

**Supplementary Figure S2.**
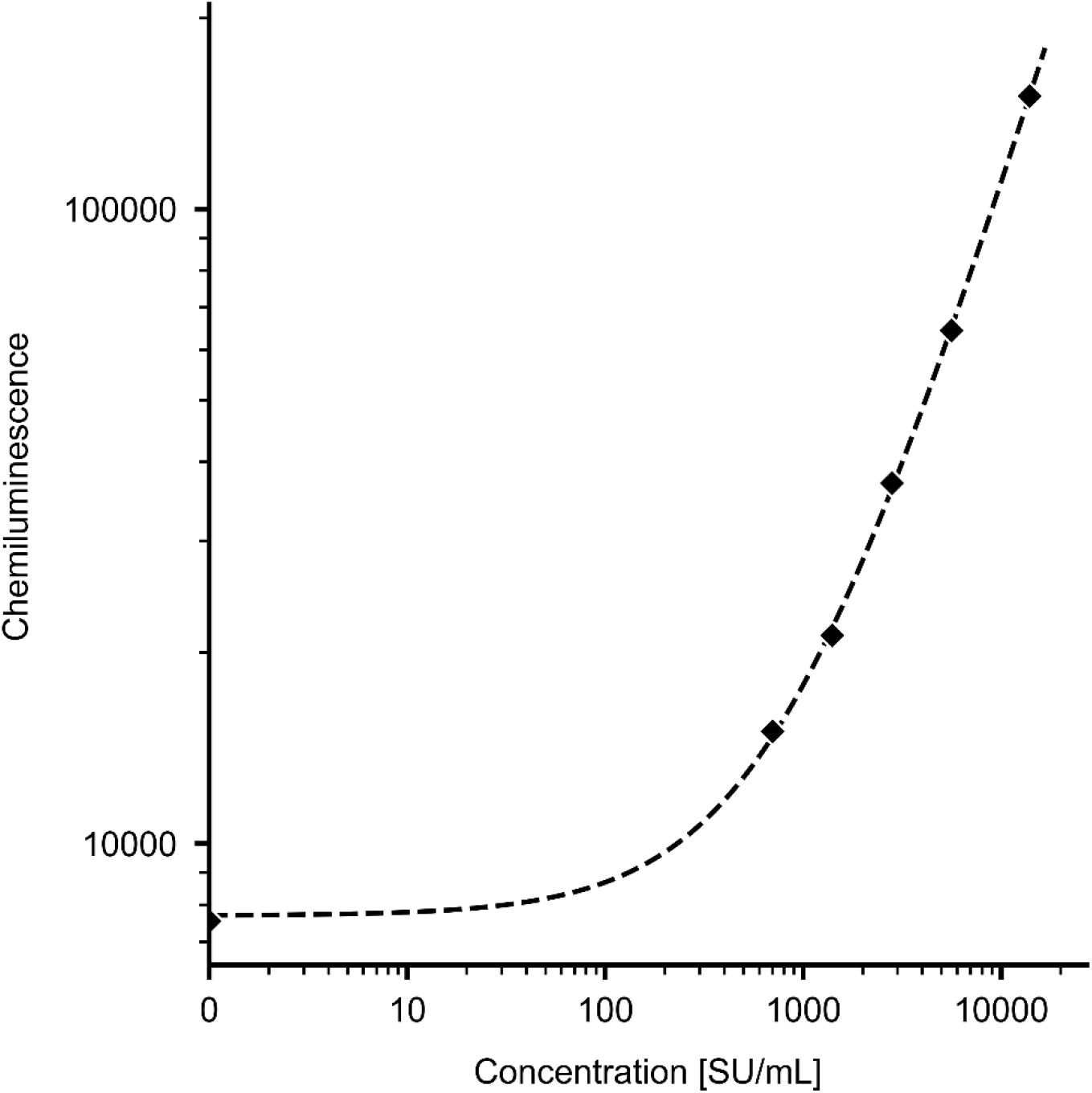
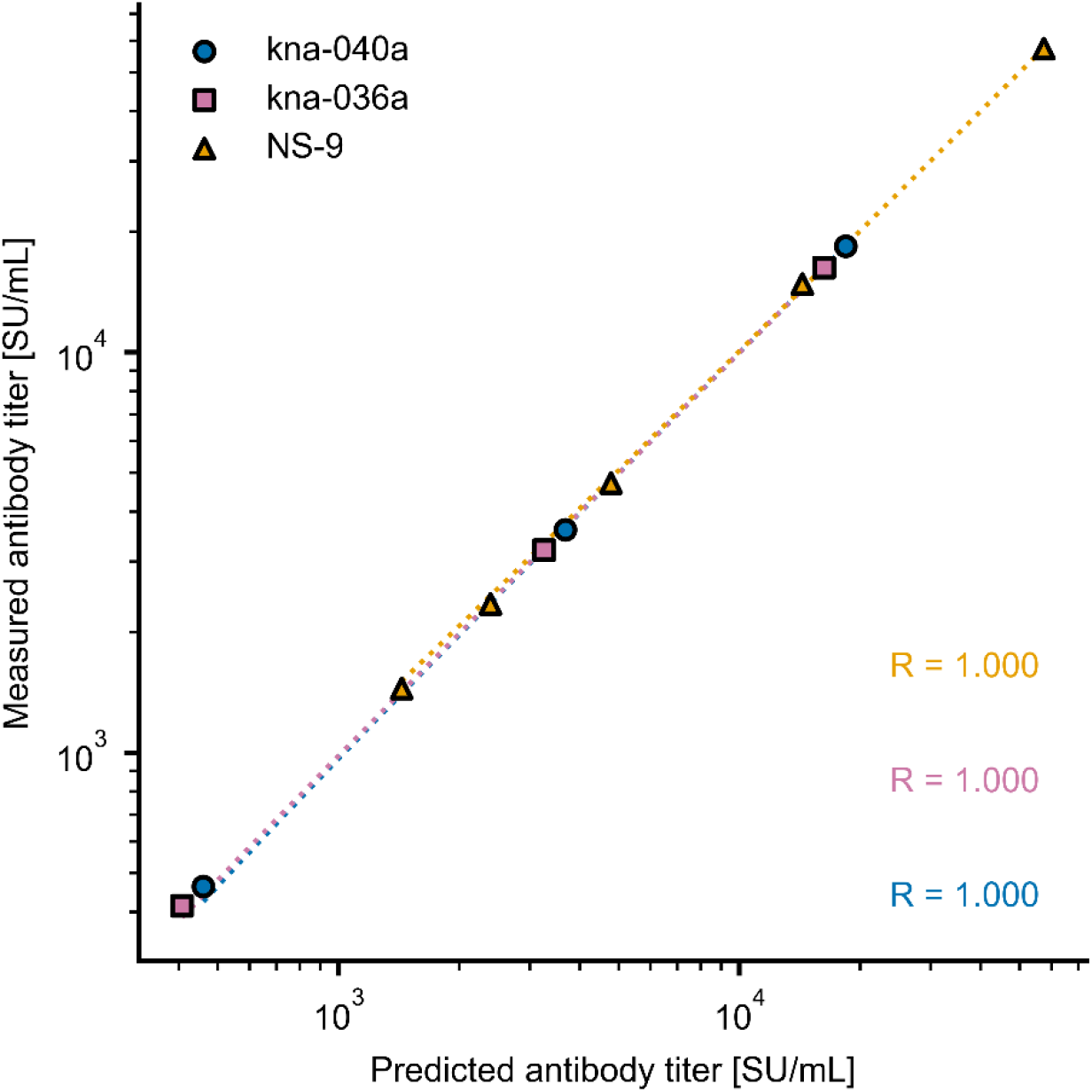
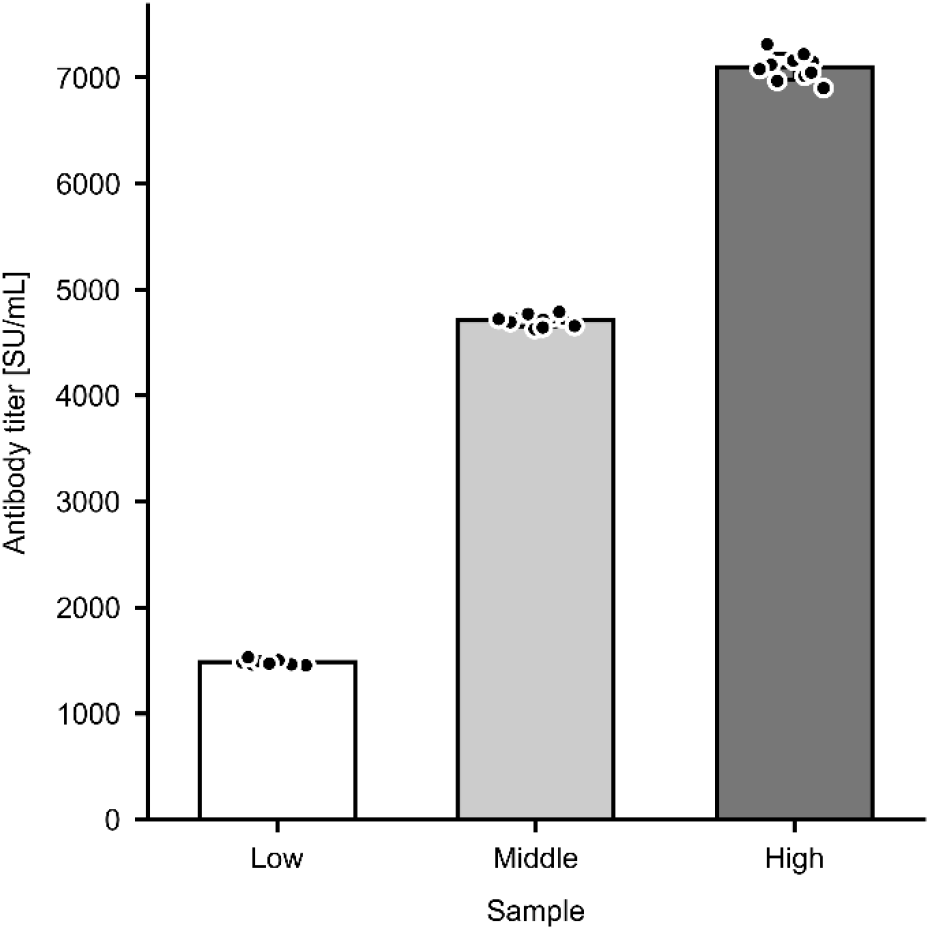
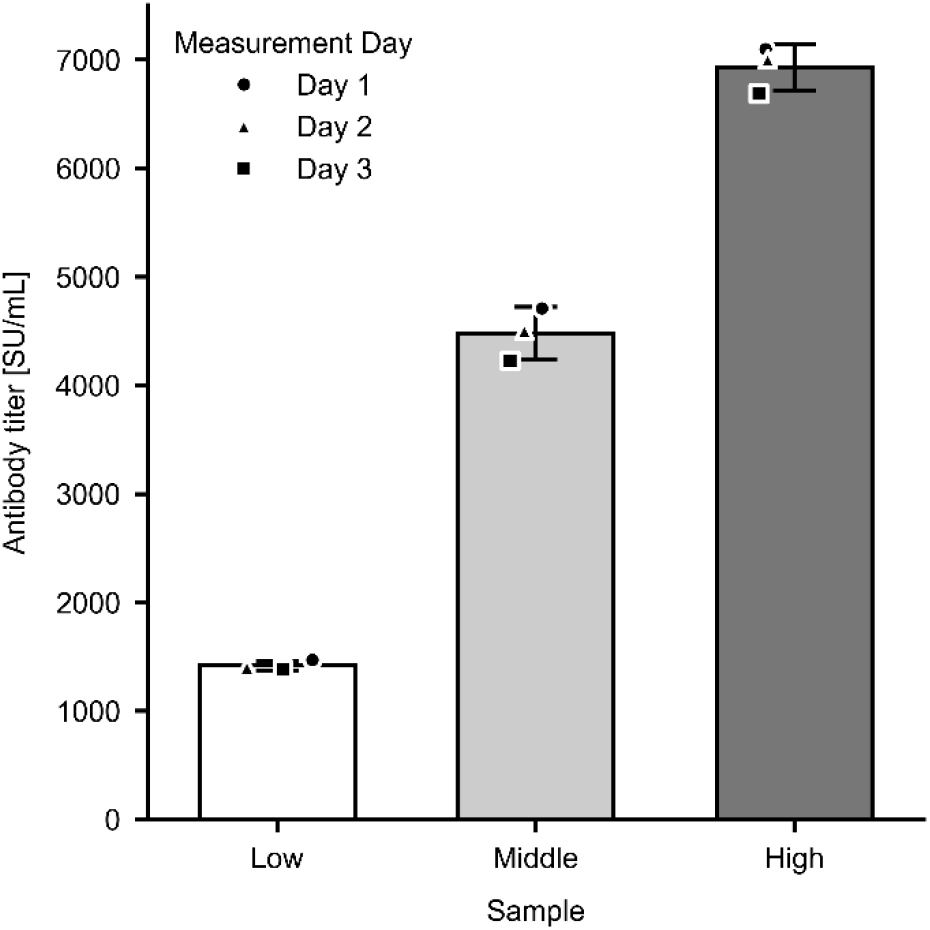
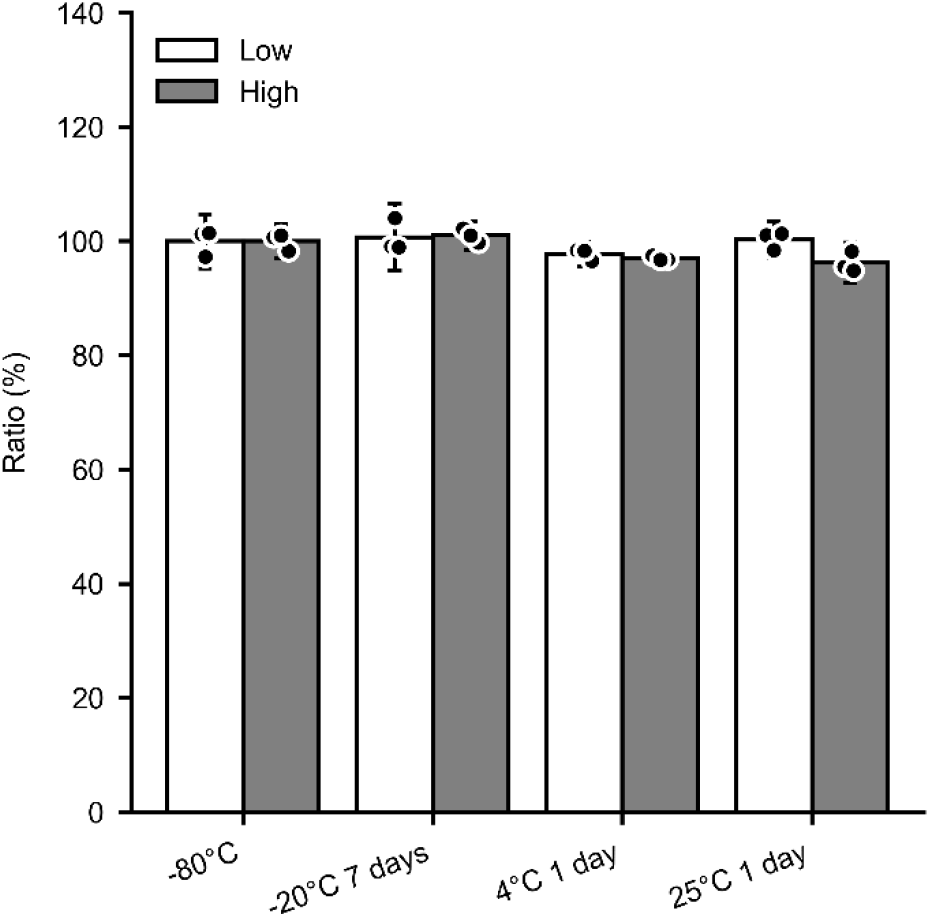
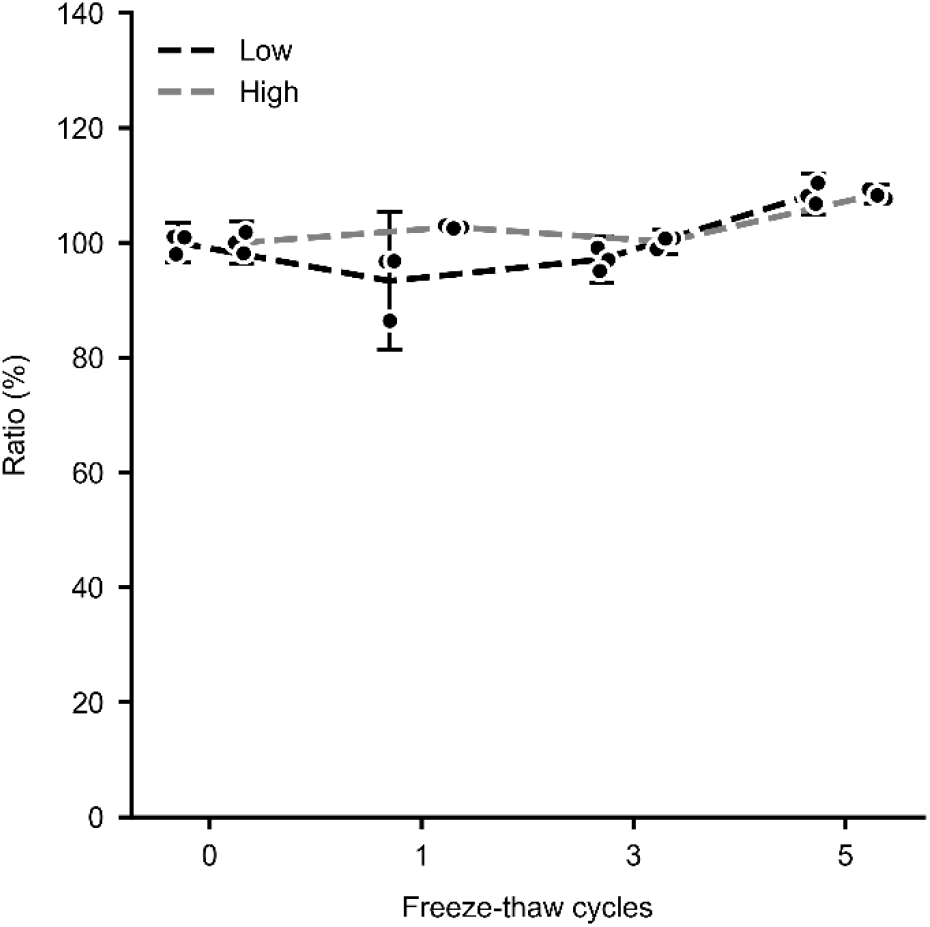
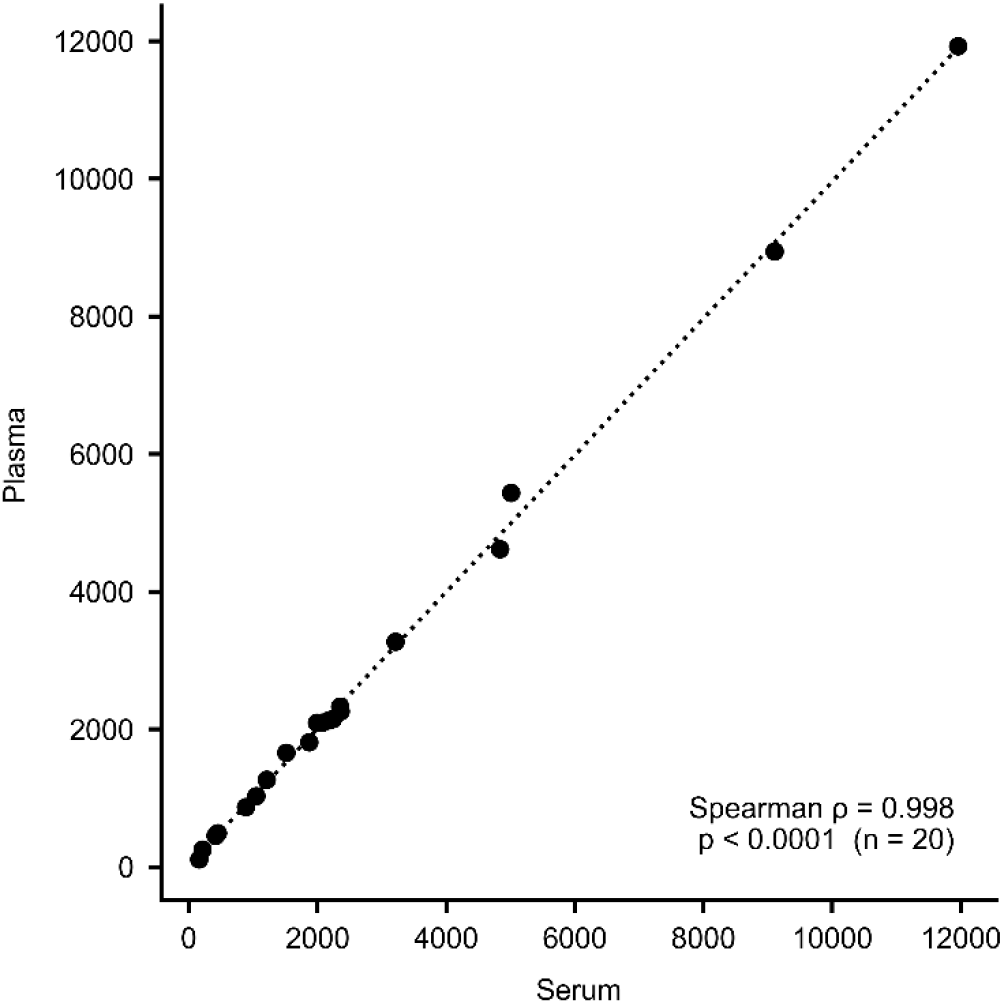
Analytical performance of the HISCL-based assay (a) Construction of the calibration curve. Calibration curve constructed using a four-parameter logistic regression model to convert chemiluminescence intensity (HISCL counts) into the assay-specific unit (SU/mL). The calibrators were prepared by serially diluting a high-titer positive sample by weight. For visualization purposes on a logarithmic scale, the blank sample (0 SU/mL) is plotted at 1 SU/mL. **(b) Evaluation of dilution linearity** Dilution linearity evaluated using three high-titer serum samples from pediatric patients with nephrotic syndrome to determine the upper limit of quantitation (ULoQ). **(c) Intra-assay reproducibility** Three representative serum samples with low (Low), middle (Middle), and high (High) anti-nephrin autoantibody concentrations were measured in 10 replicates within a single run (n = 10) for intra-assay evaluation. Data are presented as mean ± SD. **(d) Inter-assay reproducibility** Three representative serum samples with low (Low), middle (Middle), and high (High) anti-nephrin autoantibody concentrations were measured once a day over three days (n = 3) for inter-assay evaluation. Data are presented as mean ± SD. **(e) Sample stability under different storage conditions** Aliquots of two samples (low and high antibody concentrations) were evaluated across different storage conditions including -80°C (baseline control), -20°C for 7 days, 4°C for 1 day, and 25°C for 1 day. Measurements evaluations were performed in triplicate (n = 3). Data are presented as mean ± SD. **(f) Sample stability after repeated freeze-thaw cycles** Aliquots of the two samples were evaluated after up to five freeze-thaw cycles. Measurements for evaluations were performed in triplicate (n = 3). Data are presented as mean ± SD. **(g) Comparability of measurement values between serum and plasma samples** Paired serum and plasma samples from 20 pediatric patients with nephrotic syndrome (14 in the active phase and 6 in remission) were analyzed using the optimized protocol incorporating a 0.22-μm filtration step. The measurement values are shown as HISCL count, and each data point represents the mean of duplicate measurements. The dotted line indicates the regression line. Correlation was evaluated using Spearman’s rank correlation coefficient (Spearman’s ρ = 0.998, P < 0.0001).

**Supplementary Figure S3.**
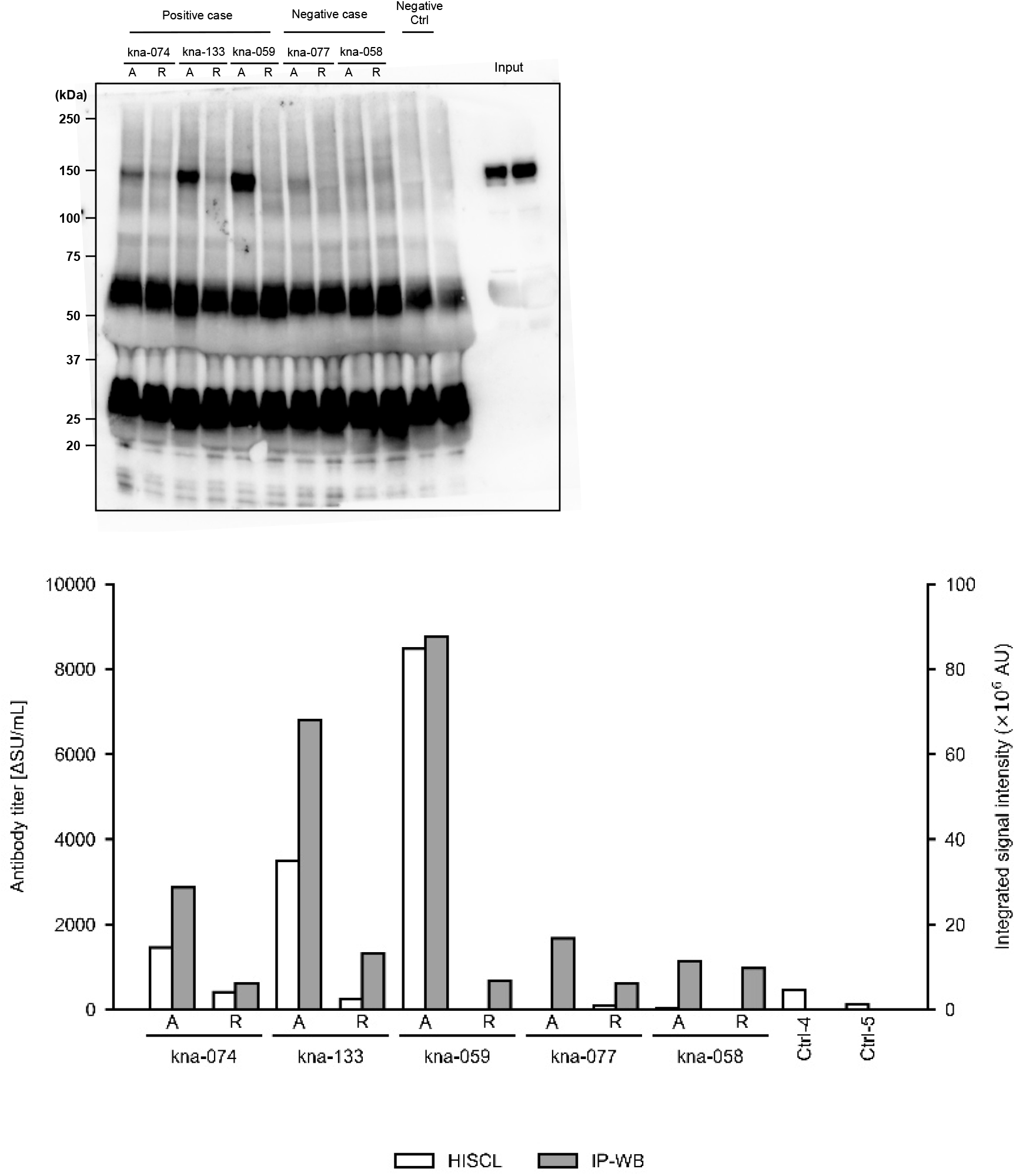
Validation of the automated immunoassay for anti-nephrin autoantibodies by immunoprecipitation–western blotting. Serum samples from pediatric patients with idiopathic nephrotic syndrome were obtained at disease onset (A) and remission (R). Healthy adult controls (Ctrl) were included as negative controls. Upper panel: representative immunoprecipitation–western blotting (IP-WB) images showing immunoprecipitated recombinant nephrin. Samples positive for anti-nephrin autoantibodies by the automated immunoassay demonstrated corresponding nephrin-specific bands by IP-WB, whereas negative samples showed no detectable reactivity. Lower panel: comparison of anti-nephrin autoantibody levels measured by the HISCL assay (left y-axis) and corresponding IP-WB signal intensities quantified by densitometric analysis (right y-axis).

**Supplementary Figure S4.**
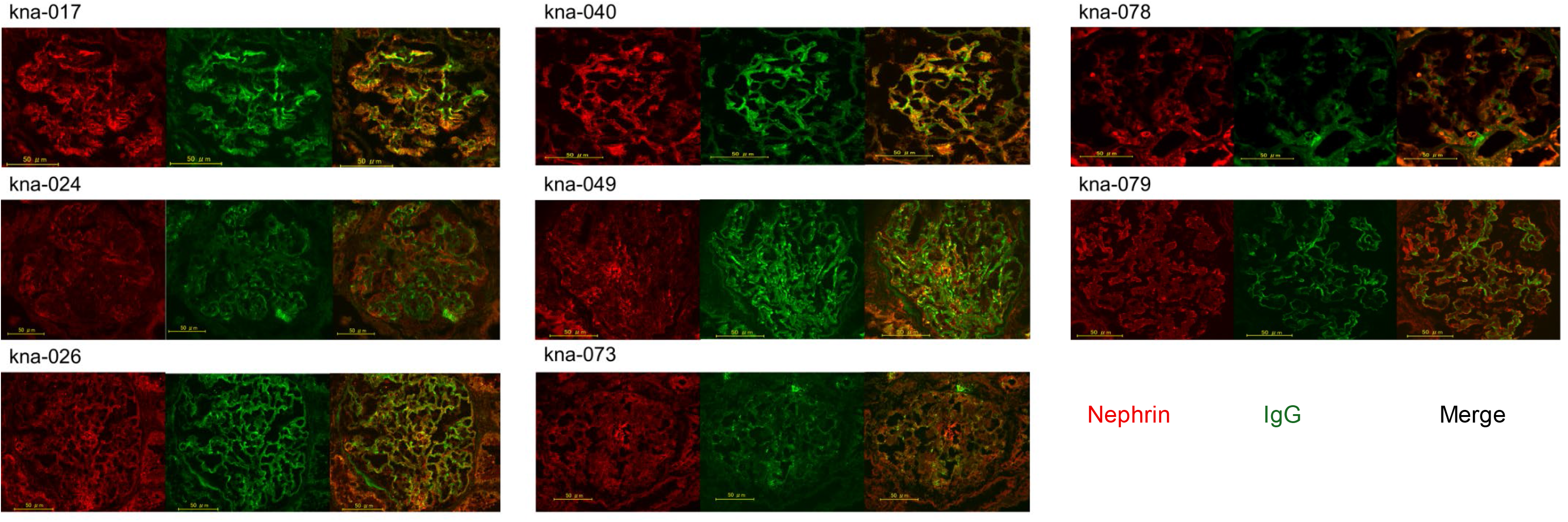
Evaluation of glomerular IgG–nephrin colocalization in kidney biopsy specimens. Double-immunofluorescence staining for nephrin (red) and IgG (green) was performed in kidney biopsy specimens from patients with available renal tissue. Merged images are shown in the right panels, with yellow fluorescence indicating colocalization of nephrin and IgG. Glomerular IgG-nephrin colocalization was detected in patients kna-017, kna-040, kna-049, and kna-073, whereas no colocalization was observed in kna-024, kna-078, and kna-079. In kna-026, small uniform IgG granules were scattered within the glomeruli and showed only partial overlap with nephrin staining, precluding definitive classification as either positive or negative for glomerular IgG-nephrin colocalization. The presence or absence of glomerular IgG–nephrin colocalization was evaluated and compared with circulating anti-nephrin autoantibody status. Scale bars, 50 μm.

**Supplementary Figure S5.**
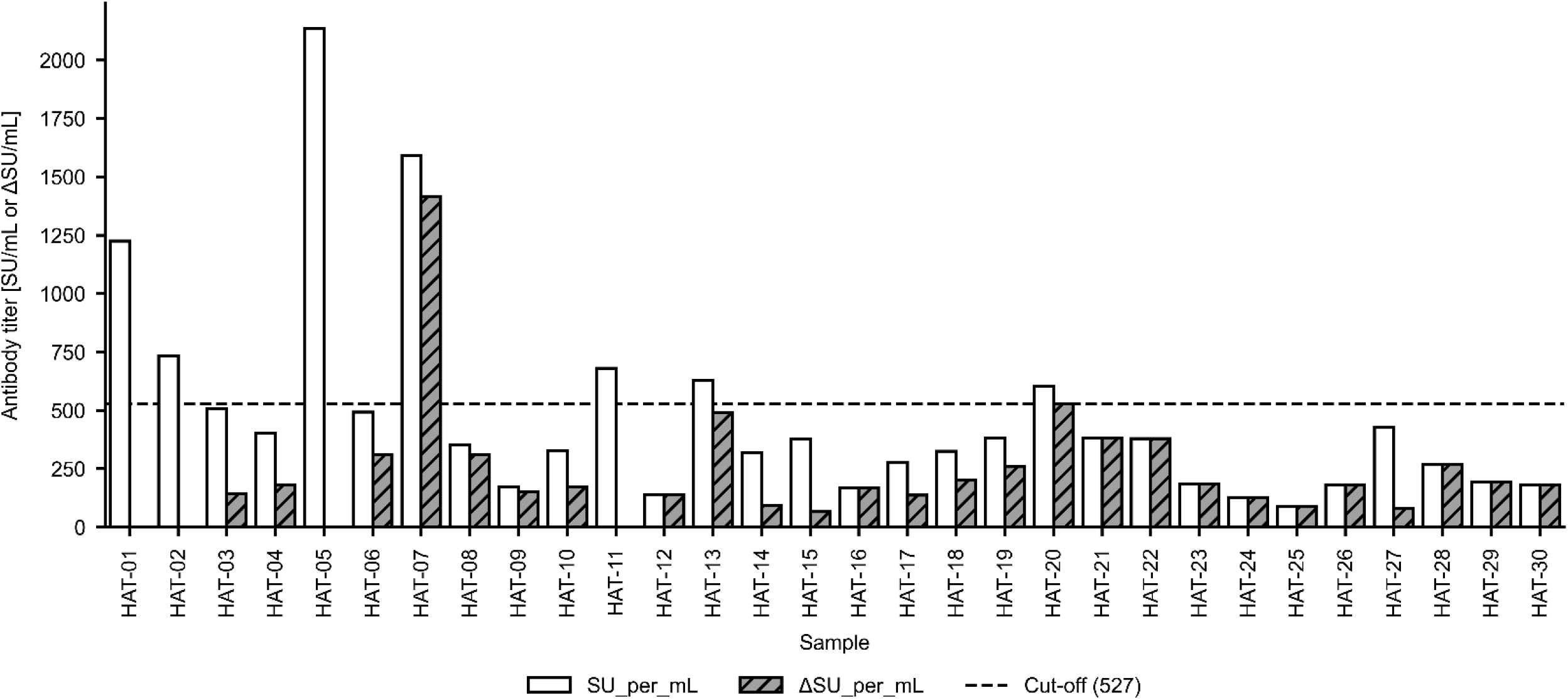
Establishment of the cut-off value for anti-nephrin autoantibody positivity. Anti-nephrin autoantibody levels were evaluated in 30 pediatric acute-phase disease control samples. The graph displays two values for each sample: the initial measurement (SU/mL) and the background-corrected measurement (ΔSU/mL). To eliminate non-specific background signals, presumably arising from the non-specific adsorption of IgG, the signal measured using bovine serum albumin (BSA)-coated magnetic particles was subtracted from the signal obtained using nephrin antigen-coated magnetic particles to determine the ΔSU/mL value. After excluding a single statistical outlier identified using Tukey’s fences method, the highest recorded value among the remaining 29 control samples (527 ΔSU/mL) was established as the threshold for positivity. The horizontal dashed line indicates this cut-off value of 527. SU, unique units.

**Supplementary Figure S6.**
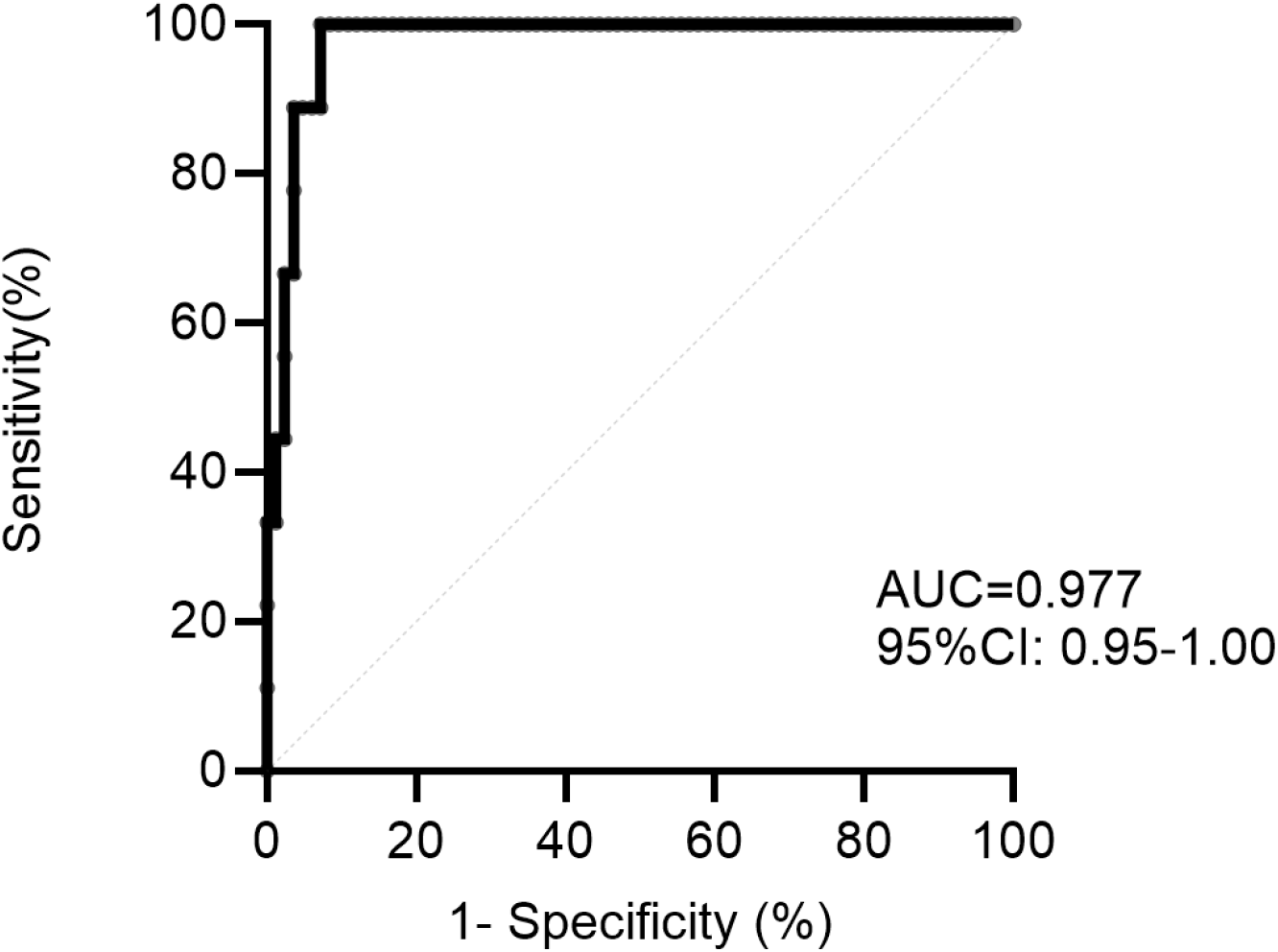
ROC curve analysis of anti-nephrin autoantibody titers for predicting steroid-resistant nephrotic syndrome. ROC curve analysis was performed using anti-nephrin autoantibody-positive patients with SSNS (n = 83) and SRNS (n = 9). The area under the curve was 0.977 (95% CI, 0.95–1.00). The optimal cut-off value determined by the maximum Youden index was 7,783 ΔSU/mL, yielding a sensitivity of 100% and a specificity of 92.77% for identifying SSNS among antibody-positive patients. . CI, confidence interval.

**Supplementary Figure S7.**
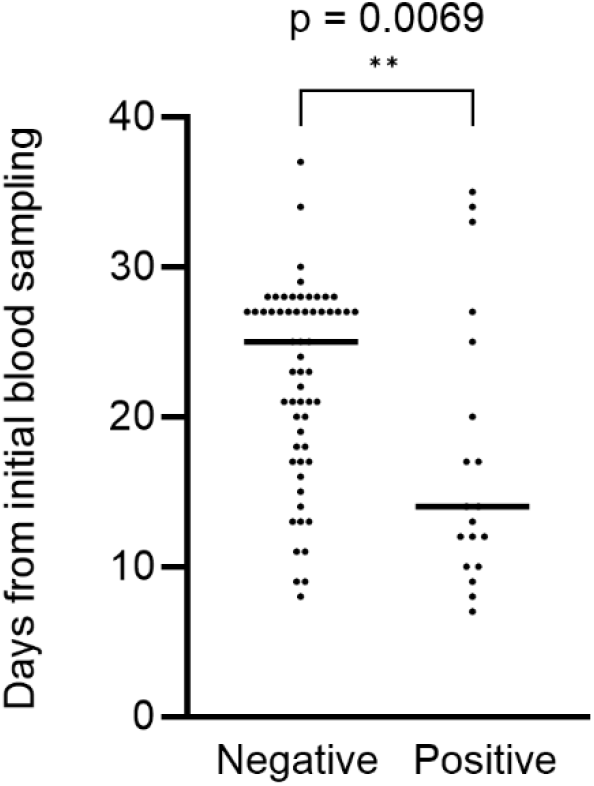
Interval from initial blood sampling to remission-phase sampling according to anti-nephrin autoantibody status at remission. Among patients with remission-phase samples available, those who remained positive for anti-nephrin antibodies during remission had a significantly shorter interval between the initial blood sampling and remission-phase sampling than those who were antibody-negative during remission (median, 14 vs. 25 days; exact Mann–Whitney test, *P* = 0.0069). Horizontal bars indicate the median values. Each dot represents an individual patient.

**Supplementary Figure S8.**
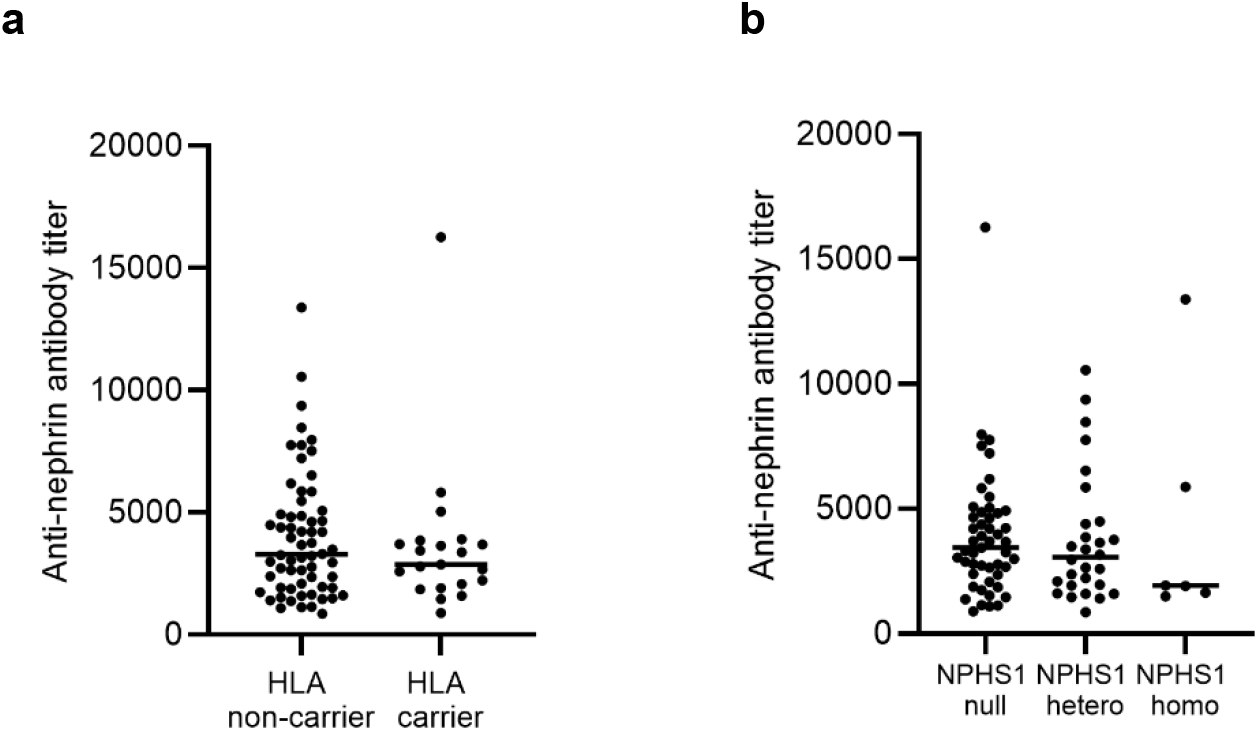
Baseline anti-nephrin autoantibody titers according to *HLA* and *NPHS1* risk allele status in SSNS. **(a)** Baseline anti-nephrin autoantibody titers in *HLA* risk allele carriers and non-carriers among patients with SSNS. No significant difference was observed (Mann–Whitney U test, *P* = 0.38; n = 83). **(b)** Baseline anti-nephrin autoantibody titers according to *NPHS1* risk allele genotype (null, heterozygous, and homozygous) in patients with SSNS. No significant difference was observed among the genotype groups (Kruskal–Wallis test, *P* = 0.7005; n = 83). Each dot represents an individual patient, and horizontal bars indicate the median and interquartile range.

**Supplementary Figure S9.**
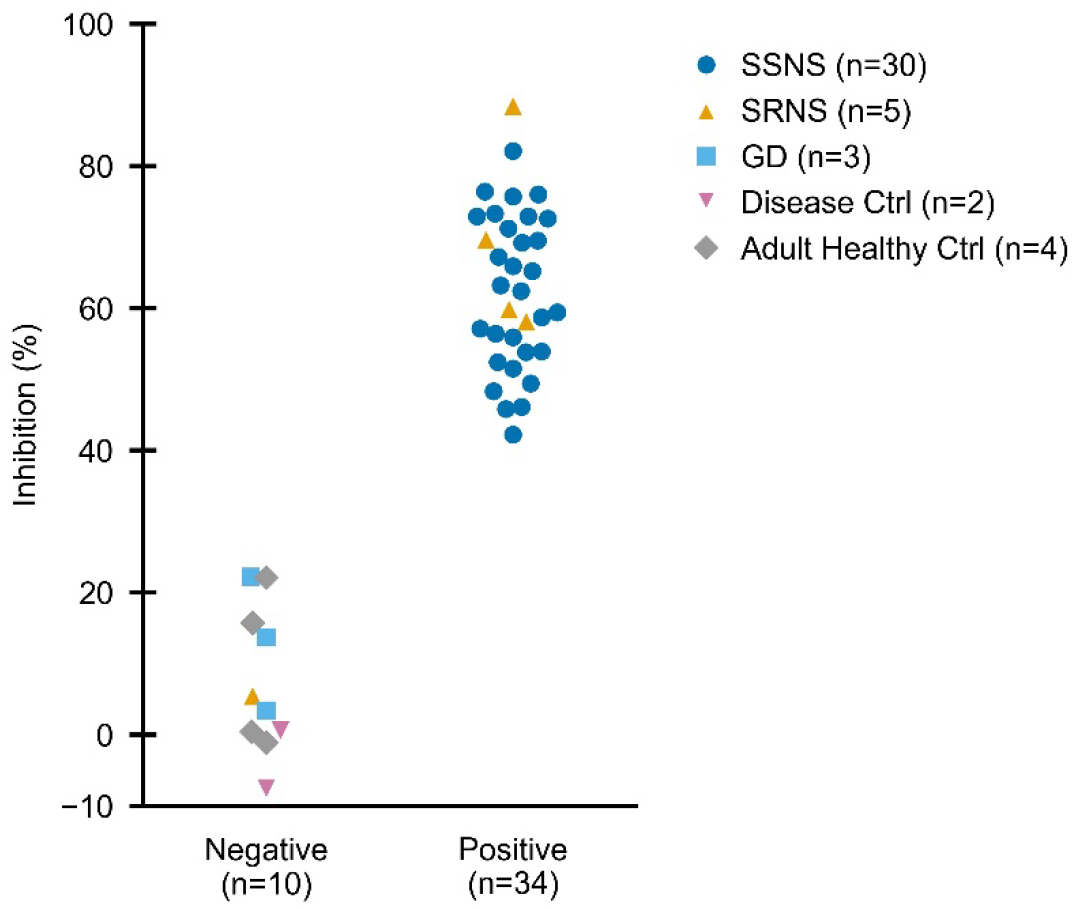
Antigen inhibition assay in the validation cohort. Inhibition rates (%) following the addition of recombinant nephrin antigen to the samples are shown for the anti-nephrin autoantibody-negative (n = 10) and -positive (n = 34) groups. Individual patients are represented by specific marker shapes corresponding to their categories: SSNS by circles (n = 30), SRNS by upward triangles (n = 5), GD by squares (n = 3), Disease Ctrl by downward triangles (n = 2), and Adult Healthy Ctrl by diamonds (n = 4).

## Japanese pediatric anti-nephrin autoantibody research consortium

Tomoko Horinouchi, Kazumoto Iijima, Kandai Nozu, Tomohiko Yamamura, China Nagano, Nana Sakakibara, Yuta Ichikawa, Yuta Inoki, Shingo Ishimori, Shuhei Aoyama, Yuka Kimura, Ayako Fujiwara, Minato Baba, Ryo Nishimura, Asahi Yamamoto, Hiroshi Kaito, Yu Tanaka, Ryota Suzuki, Takayuki Okamoto, Hiroki Miyano, Kyosuke Tabata, Koichi Nakanishi, Wataru Shimabukuro, Shogo Nakada, Tomoki Akamine, Erika Hidawa, Moe Yoshimura, Yasuyo Kashiwagi, Yoshimitsu Gotoh, Hisakazu Majima, Toshihiko Hattori, Yuki Ashida, Miwa Goto, Anna Kobayashi, Yuko Hamasaki, Ayuko Zaitsu, Koichi Kamei, Masao Ogura, Kentaro Nishi, Yohei Ikezumi, Tomomi Kondoh, Takuma Ando, Eriko Tanaka, Taishi Nada, Yu Kamigaki, Toru Uchimura, Riku Hamada, Kimura Naoki, Yusuke Iizuka, Natsumi Takeshi, Hitoshi Nakazato, Hiroshi Tamura, Natsumi Yamamura, Kaori Fujiwara, Shinya Ishiko, Junya Fujimura, Kenji Ishikura, Yusuke Okuda, Chikako Terano, Shinya Kon, Kaori Kikunaga, Yoshinori Araki, Azusa Kawaguchi, Rika Fujimaru, Tomoo Kise, Masatsugu Uehara, Yoshitsugu Kaku, Rie Sato, Yuka Watanabe, Yutaka Harita, Yoko Shirai, Kenichiro Miura, Naoya Fujita, Kazuki Tanaka, Mayuko Terasawa, Hidehito Kondo, Masatoshi Yoshikane, Hideki Ban, Yasuko Kobayashi, Chikage Yajima, Yoko Yamasaki, Yuko Shima, Takaya Hasegawa, Naohiro Kamiyoshi, Shoji Tsuji, Shunsuke Shinozuka, Yoshiharu Nagaoka, Masayuki Ishihara, Rie Kuroki, Tae Omori, Yuya Aoto, Takeshi Ninchoji, Tamaki Morohashi

